# Estimating the risk reduction of isolation on COVID-19 non-household transmission and severe/critical illness in non-immune individuals: September to November 2021

**DOI:** 10.1101/2022.02.05.22270453

**Authors:** Aaron Prosser, Bartosz Helfer, David L. Streiner

**Affiliations:** Department of Psychiatry and Behavioural Neurosciences, McMaster University, Hamilton, Ontario, Canada; Meta-Research Centre, Institute of Psychology, University of Wroclaw, Wroclaw, Lower Silesian Voivodeship, Poland; National Heart and Lung Institute, Imperial College London, London, UK

**Keywords:** SARS-CoV-2, COVID-19, transmission, severe illness, critical illness

## Abstract

In the fall 2021, immunity mandates/passports for COVID-19 started to be discussed and implemented globally. In addition to increasing vaccination levels, these interventions isolate non-immune individuals from various settings to reduce non-household transmission and severe/critical illness. This is based on the hypothesis that the non-immune are at high absolute risk of these outcomes. However, these absolute risks were not quantified in the literature such that the absolute risk reductions of isolation on these outcomes remain unknown. This study estimated these absolute risks from September to November 2021 prior to the emergence of Omicron (B.1.1.529) using known data on the risk of infection, transmission in non-household settings, and age-stratified severe/critical illness in non-immune individuals for the Delta (B.1.617.2) variant, focusing on the European Union, United Kingdom, United States, Canada, Australia, and Israel. This allowed us to quantify the absolute risk reductions of isolation on (1) non-household transmission from the non-immune and (2) severe/critical illness amongst the non-immune in these regions during this period. We observed that on any given day the absolute risk reductions of isolation were typically small for transmission in most types of non-household settings and severe/critical illness in most age-groups, especially those aged <40. During a wave or sustained higher infection risks, the risk reductions were modest only for transmission in intimate social gatherings and severe/critical illness in adults aged ≥50-60. The limitations of this study and the implications for the expected benefits of isolating non-immune individuals on reducing these outcomes are discussed.

## Introduction

In the fall 2021, immunity mandates/passports (IMP) for SARS-CoV-2 started to be discussed/implemented in many countries, notably the European Union (EU), United Kingdom (UK), United States (US), Canada, Australia, and Israel. In addition to increasing vaccination, IMP isolate those who remain non-immune in order to (i) reduce non-household transmissions from the non-immune and (ii) reduce severe/critical illness amongst the non-immune. These benefits are based on the hypothesis that the non-immune are at high absolute risk (AR) of these outcomes, thereby warranting isolation. IMP isolate non-immune individuals from various settings, thus limiting their (i) contact with others and (ii) exposures and, in turn, their risk of developing a severe/critical infection. However, these ARs were not quantified in the scientific literature, such that the absolute risk reductions (ARRs) of isolation during the fall 2021 remain unknown. In essence, these ARs are the ARRs of IMP on these outcomes because isolating non-immune individuals removes these risks from the general population. Therefore, like the number needed to treat (NNT=1/ARR) [1], one can quantify the risk reductions gained using isolation by taking the reciprocal of these ARs to convert these probabilities into a more intuitive form (“1 in X”). This becomes what can be called the ‘number needed to isolate’ (NNI), which is the number of non-immune individuals needed to isolate to prevent one transmission event or one case of severe/critical illness. This paper calculated the NNIs during this period to quantify the risk reductions of isolation, which was a time when Delta (B.1.617.2) predominated. The period from September 1 to November 26, 2021, was studied because the latter date was when Omicron (B.1.1.529) was declared a variant of concern (VOC). Shortly afterwards a new phase of the pandemic started where Omicron predominated. Therefore, the fall period prior to this date was the period of interest.

## Methods

Estimating the NNIs for these outcomes requires estimating (i) the AR of a transmission event in non-household settings (*AR*_*tr*_) and (ii) the AR of severe (*AR*_*sv*_) or critical (*AR*_*cr*_) illness for the Delta variant in non-immune individuals. *AR*_*tr*_ is the probability of a transmission event in a non-household setting from a non-immune person in the general population infected with the Delta variant. This risk is estimated by taking the combined probability of the risk of infection (IR) and the risk of transmission from a non-immune person in that type of non-household setting (e.g., healthcare). The latter is the secondary attack rate (SAR) of a Delta infection typically observed from non-immune index cases in that type of setting:

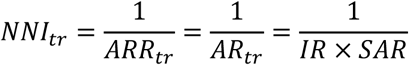

*ARR*_*tr*_ is the absolute risk reduction of isolation on transmission from non-immune people in a given type of non-household setting. The combined probability is needed to estimate *AR*_*tr*_ because a person must be infected first before they can transmit SARS-CoV-2. Technically, this *AR*_*tr*_ is the risk of one transmission *event*, which may include one or more secondary infections. This is because the SAR is the proportion of infections amongst the contacts of an index case, such that the total number of secondary infections depends on the total number of contacts. For example, a SAR of 20% is consistent with 20/100, 2/10, and 1/5. *AR*_*tr*_ is the risk of one generation of transmission caused by the non-immune index case, assuming they go into a setting of that type while infected. The IR is the point-prevalence of infectious cases in the general population, which is the estimated risk that a non-immune individual is infected.

*AR*_*sv*_ and *AR*_*cr*_ are the probabilities that a non-immune person in the general population gets a Delta infection which develops into a severe or critical illness, respectively. Given the steep age-risk gradient for severe/critical illness from SARS-CoV-2 [2], it is important to stratify these ARs by age. These ARs are estimated by taking the combined probability of the IR and age-stratified rates of severe illness (infection-severe rates, ISRs) and rates of critical illness (infection-critical rates, ICRs) amongst non-immune individuals infected with the Delta variant. The combined probability is needed because one cannot develop a severe/critical illness unless one is first infected with SARS-CoV-2. Therefore the NNI for severe illness is estimated:

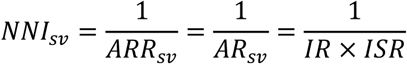

and similarly for critical illness:

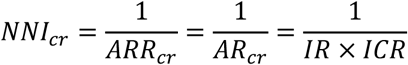

*ARR*_*sv*_ and *ARR*_*cr*_ are the absolute risk reductions of isolation on severe and critical illness, respectively, amongst non-immune people in a given age-group.

Like the NNT, time is implicit in the NNI since it relates to the time window over which the risk was measured. These ARs are the risk on a given day (i.e., the day of the IR) because point-prevalence data are typically measured over one day. Moreover, the contact duration in most non-household settings is typically less than one day. For these reasons, the NNI is the number of non-immune individuals needed to isolate *on that day* (i.e., the day of the IR) to prevent one transmission event or one severe/critical illness. In order to show the risk reductions of isolation during a time period, NNIs can be calculated over time using daily IRs, which is what we did in this paper. This is an important difference between the NNI and NNT. For the NNI, the ARR is based on point-prevalence and the time window is one day. For the NNT, the ARR is typically based on incidence proportion and the time window is often months or years. The rationale for using the point-prevalence rather than other metrics (e.g., incidence, period prevalence, forecasted risks) to estimate these ARs is detailed in the Discussion.

Daily IR point-estimates and 95% confidence intervals (CIs) from September 1 to November 26, 2021, inclusive were taken from the Defence Research and Development Canada (DRDC) database [3]. Its estimation methods are detailed online. The daily point-prevalence of infectious cases from this period were extracted for the EU member states, US states, Canadian provinces/territories, Australian states and the Capital/Northern Territory, and Israel. US county data were available and means were used to calculate state-level daily IRs. Data were extracted on January 25 to 28, 2022. The daily IR point-estimates in each region were used to calculate the NNIs on each day for non-household transmission and severe/critical illness. Box and whisker plots were used to display the distribution of the NNIs during this period in each region to show the ARRs of isolating non-immune individuals on these outcomes.

As noted, these IRs must be multiplied by estimates of the non-household SARs, ISRs, and ICRs of non-immune individuals infected with the Delta variant. Almost all the transmission data for the Delta variant involve households, which means the SARs of non-household settings must be estimated. This can be done using known data on the non-household SARs of the wild-type. There is a literature on these wild-type SARs since this was an area of focus during 2020. The COVID-19 vaccines were not available during this period. There was also relatively low natural immunity, as shown by the global median seroprevalence of SARS-CoV-2 antibodies in the general population in 2020 (median 4.5%, IQR: 2.4%, 8.4%) [4]. In other words, this data is ideal for our purposes because it captures the SARs of non-immune index cases in different types of non-household settings. Therefore, the Delta SAR in a given type of non-household setting (*SAR*_*Delta*_) can be estimated by multiplying the wild-type SARs (*SAR*_*wt*_) by a correction factor (*CF*_*Delta*_=1.97) to account for the increased transmissibility of Delta over the wild-type, which is about 97% more transmissible based on a global analysis of reproduction numbers [5]:

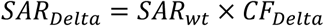

A systematic search identified 7 meta-analyses of the wild-type SARs [6-12] (see Supplemental Appendix). Delta SARs in six types of settings were estimated after applying this procedure to the mean SARs across these meta-analyses: households (mean SAR=32.59%), social gatherings (mean SAR=11.69%), casual close contacts (mean SAR=3.05%), work/study places (mean SAR=2.89%), healthcare (mean SAR=2.96%), and travel/transportation (mean SAR=4.40%) (Table S1). The *AR*_*tr*_ for each non-household setting was estimated by multiplying its mean SAR by the IRs. Social gatherings are intimate settings where the intensity of contact is less than households but still high (e.g., gatherings of friends/family), whereas casual close contacts are lower intensity contacts (e.g., public areas/buildings). Notably, the estimated household SAR (32.59%) matched the observed mean SAR in a meta-analysis of household transmission of the Delta variant (30.8%) [13]. This suggests this procedure likely produced accurate estimates of the non-household SARs for Delta infections.

A similar procedure was needed for the ISRs and ICRs. The first and only meta-analysis of seroprevalence studies to calculate age-stratified ISRs and ICRs for SARS-CoV-2 was recently made available [14]. The ISR was defined as those resulting in hospitalisation or out-of-hospital death. The ICR was defined as those resulting in ICU admissions or out-of-ICU deaths. The data were from early to mid-2020 when the wild-type predominated and immunity was low. Therefore, these ISRs and ICRs likely capture the risk of severe/critical illness in non-immune people. For this reason we extracted the mean ISR and ICR in each age-group in this report to estimate *AR*_*sv*_ and *AR*_*cr*_. Age-stratified correction factors (*CF*_*Delta*_) are needed to account for the increased severity of Delta vs. the wild-type. For this we used the significant adjusted odds ratios for hospitalisation and ICU admission, respectively, of Delta vs. wild-type infections from a large retrospective cohort (Table S2) [15]. The age-stratified ISR for the Delta variant was estimated:

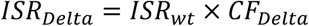

and similarly for critical illness:

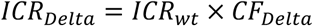

## Results

From September to November 2021, IRs on any given day were typically ≤ 5% and stable (Figure 1) because there was no wave of infection in most regions, specifically the UK, many western, southern, and northern EU countries (Figure S1), Israel (Figure S2), many parts of Canada (Figure S3), Australia (Figure S4), and many states of the US Northeast (Figure S5), Midwest (Figure S6), South (Figure S7), and West (Figure S8). In central, eastern, and Balkan EU states, there was a wave (IRs=5-10%) in November prior to Omicron’s rise (Figure S1). In other EU states (e.g., Romania) an October wave subsided into November. In Canada, a wave (IRs=5-10%) in September in Alberta, Saskatchewan, and the Northwest Territories gradually subsided into October, whereas the Yukon saw a wave in November (Figure S3). In some parts of the US Northeast, there was a slight upward trend in IRs (∼5-6%) into November (Figure S5). In the US Midwest, Nebraska was an outlier with a large wave in September/October which subsided in November (Figure S6). Visualization of the Nebraska data (not shown) showed the high IRs were mainly from counties with low populations (< 10,000), which may explain the high prevalence. In most of the US South, September had IRs between 5-10% which subsided by October (Figure S7). In the US West during September and October, the waves (IRs=5-15%) in Alaska, Montana, and Nevada receded by November (Figure S8).

**Figure 1.**
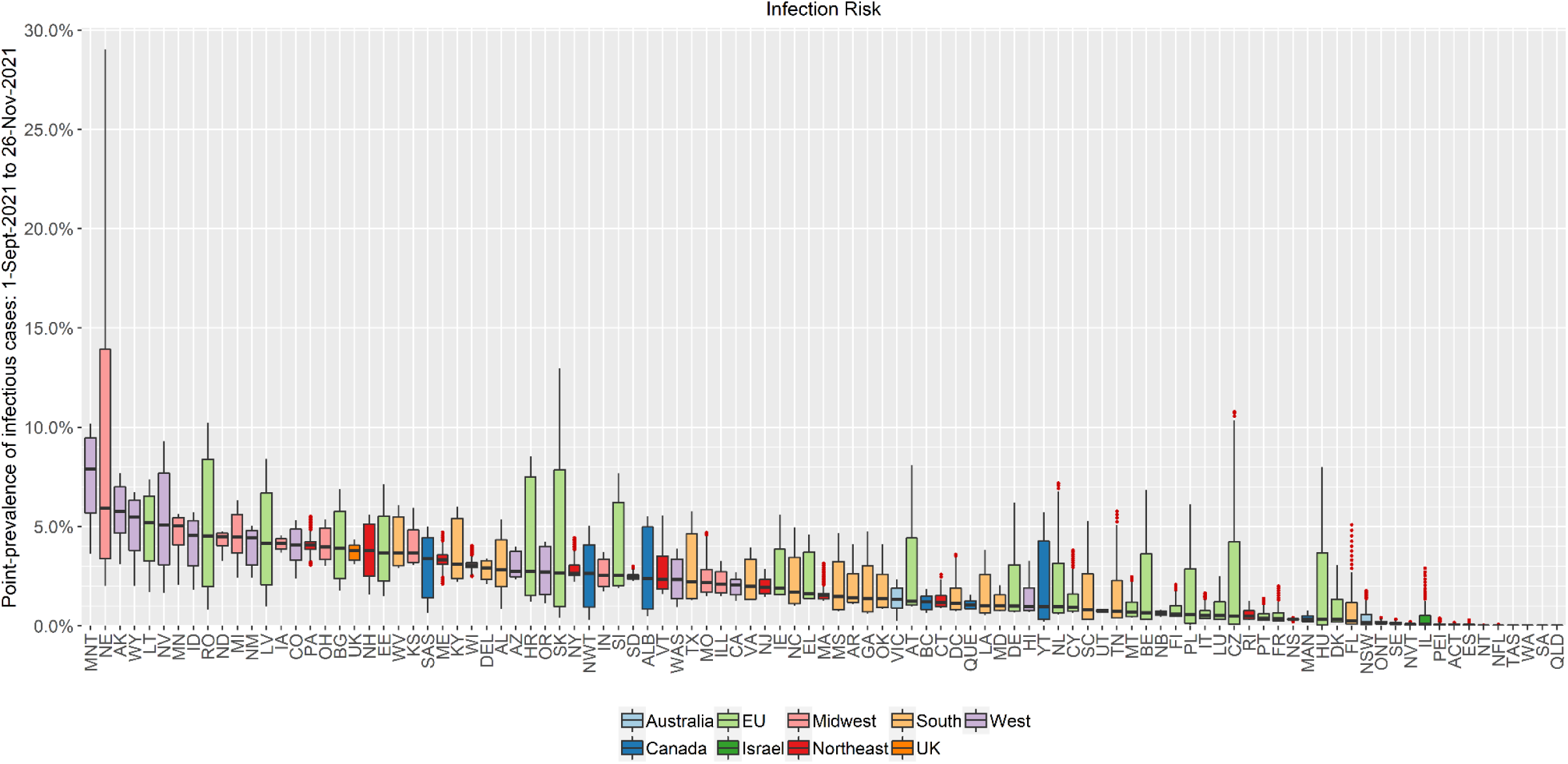
Daily infection risks from September 1 to November 26, 2021, by region and major jurisdiction. Major jurisdictions examined were EU member states, the United Kingdom (UK), Israel, Canada, Australia, and the major US regions (Northeast, Midwest, South, West). The distribution of risks during this period are displayed using box-and-whisker plots. ACT=Australian Capital Territory, NSW=New South Wales, NT=Northern Territory, QLD=Queensland, SA=South Australia, TAS=Tasmania, VIC=Victoria, WA=Western Australia, AT=Austria, BE=Belgium, BG=Bulgaria, ALB=Alberta, BC=British Columbia, MAN=Manitoba, NB=New Brunswick, NFL=Newfoundland and Labrador, NS=Nova Scotia, NVT=Nunavut, NWT=Northwest Territories, ONT=Ontario, PEI=Prince Edward Island, QUE=Quebec, SAS=Saskatchewan, YT=Yukon, HR=Croatia, CY=Cyprus, CZ=Czechia, DK=Denmark, EE=Estonia, FI=Finland, FR=France, DE=Germany, EL=Greece, HU=Hungary, IE=Ireland, IL=Israel, IT=Italy, LV=Latvia, LT=Lithuania, LU=Luxembourg, MT=Malta, NL=Netherlands, PL=Poland, PT=Portugal, RO=Romania, SK=Slovakia, SI=Slovenia, ES=Spain, SE=Sweden, UK=United Kingdom, AL=Alabama, AK=Alaska, AZ=Arizona, AR=Arkansas, CA=California, CO=Colorado, CT=Connecticut, DEL=Delaware, DC=District of Columbia, FL=Florida, GA=Georgia, HI=Hawaii, ID=Idaho, ILL=Illinois, IN=Indiana, IA=Iowa, KS=Kansas, KY=Kentucky, LA=Louisiana, ME=Maine, MD=Maryland, MA=Massachusetts, MI=Michigan, MN=Minnesota, MS=Mississippi, MO=Missouri, MNT=Montana, NE=Nebraska, NV=Nevada, NH=New Hampshire, NJ=New Jersey, NM=New Mexico, NY=New York, NC=North Carolina, ND=North Dakota, OH=Ohio, OK=Oklahoma, OR=Oregon, PA=Pennsylvania, RI=Rhode Island, SC=South Carolina, SD=South Dakota, TN=Tennessee, TX=Texas, UT=Utah, VT=Vermont, VA=Virginia, WAS=Washington, WV=West Virginia, WI=Wisconsin, WY=Wyoming.

Overall, the UK had the highest median daily IR from September to November 2021 (median 3.8%, IQR: 3.3%, 4.0%), followed by the US Midwest (median 3.5%, IQR: 2.6%, 4.5%), US West (median 3.4%, IQR: 2.1%, 5.0%), US Northeast (median 2.3%, IQR: 1.5%, 3.5%), US South (median 1.8%, IQR: 1.0%, 3.2%), EU member states (median 1.3%, IQR: 0.4%, 3.4%), Canada (median 0.5%, IQR: 0.1%, 1.3%), Israel (median 0.08%, IQR: 0.02%, 0.51%), and Australia (median 0.011%, IQR: 0.002%, 0.069%) (Figure S9). The time trend plots of the UK, US Midwest, US West, and US Northeast showed that they had the highest daily IRs mainly because the UK and many states in these regions had sustained higher IRs during this period (IRs=2-5%).

The NNIs for a non-household transmission event and a case of severe/critical illness from September to November 2021 were estimated using these IRs. The NNT can help interpret these NNIs, while recognizing that the outcomes and time windows of the NNI vs. NNT differ (see Discussion). The NNTs of acetylsalicylic acid (ASA) for primary prevention of cardiovascular disease (CVD) outcomes are ≥250, which are considered low ARRs [16]. The NNTs of ASA and statins for secondary prevention of CVD outcomes typically range between 50 and 250 [17, 18]. Graphs were cut-off at NNIs > 5,000 due to the very high NNIs for some regions/outcomes (indicating very low ARRs) which, if plotted, masked the lower end of the distribution which was important to visualize. To ease interpretation of the large amount of data we analysed, we report if the 1^st^ quartile (Q1) of the NNIs was ≥500. This threshold indicates that on ≥75% of the days from September to November 2021 in the region, the ARRs of isolation on a given day was within the range considered ‘low’.

In the EU and UK, the Q1 of the NNIs for transmission in social gathering settings was ≥500 in Cyprus, Malta, Finland, Italy, Luxembourg, Portugal, France, Denmark, Sweden, and Estonia (Figure 2). This is due to the combination of a higher SAR for social gatherings (11.69%) and higher IRs (≥5%) in the UK and other EU countries, many of whom experienced a wave during this period (Figure S1). Predictably, non-household settings with smaller SARs compared to social gatherings had higher NNIs. The NNIs were similar for these other settings because the SARs were similar: casual close contacts (3.05%), healthcare (2.96%), work/study places (2.89%), and travel/transportation (4.40%) (Table S1). Specifically, the Q1 of the NNIs for transmission in causal close contacts settings was ≥500 in the EU states and UK except for Romania, Latvia, Croatia, and Slovakia. In healthcare and work/study settings, the Q1 was ≥500 except in Romania, Croatia, and Slovakia. In travel/transportation settings, the Q1 was ≥500 except in Lithuania, Romania, Latvia, Bulgaria, Estonia, Croatia, Slovakia, and Slovenia.

**Figure 2.**
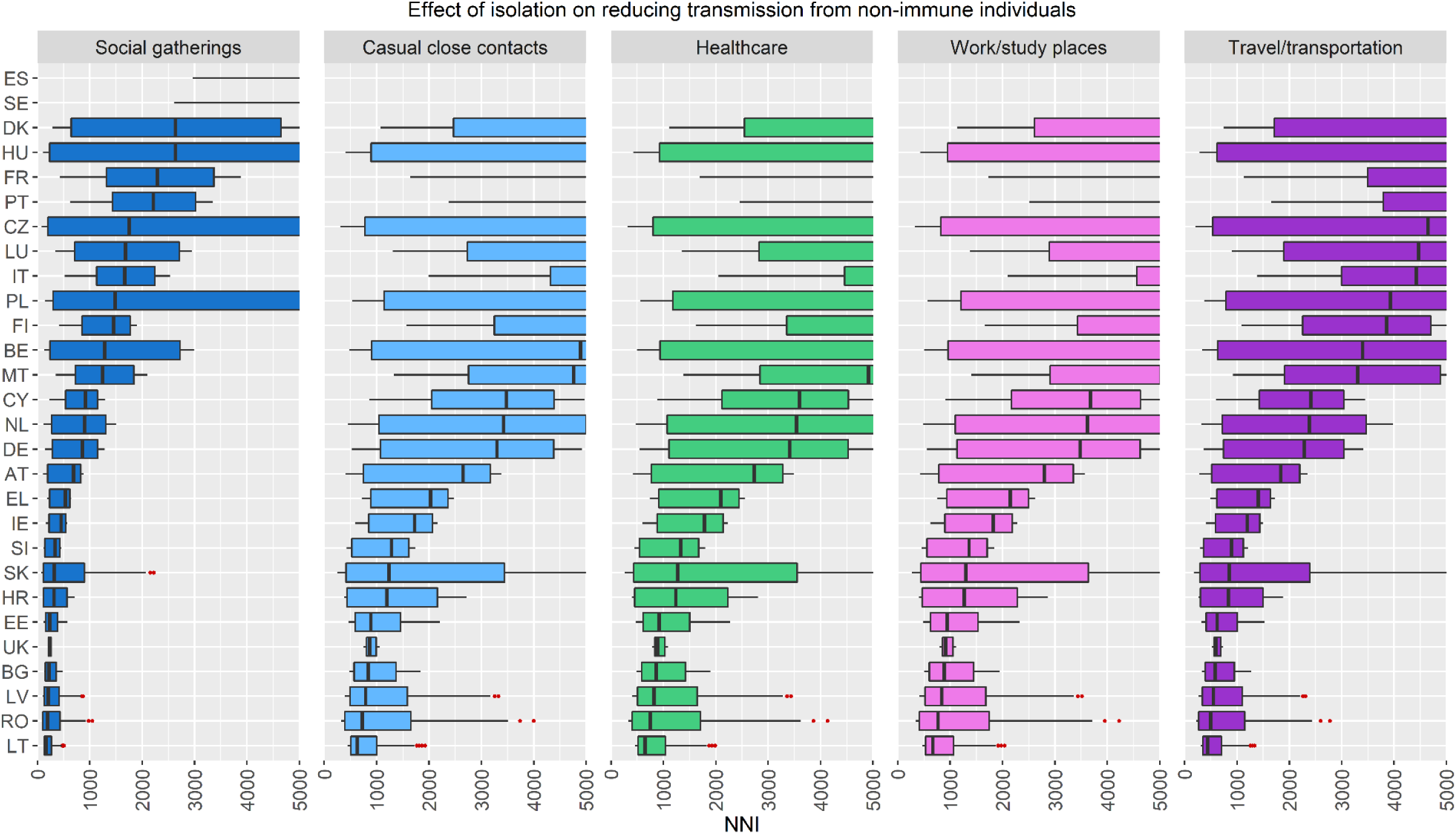
NNIs for transmission in non-household settings in the European Union member states and United Kingdom. A region/setting with no box-and-whisker plot has an NNI > 5,000 indicating a very low absolute risk reduction. AT=Austria, BE=Belgium, BG=Bulgaria, CY=Cyprus, CZ=Czechia, DE=Germany, DK=Denmark, EE=Estonia, EL=Greece, ES=Spain, FI=Finland, FR=France, HR=Croatia, HU=Hungary, IE=Ireland, IT=Italy, LT=Lithuania, LU=Luxembourg, LV=Latvia, MT=Malta, NL=Netherlands, PL=Poland, PT=Portugal, RO=Romania, SE=Sweden, SI=Slovenia, SK=Slovakia, UK=United Kingdom.

The Q1 of the NNIs for severe illness were ≥500 in the EU and UK in all age-groups under 40 (Figure 3). Between ages 40 and 60 the Q1 fell below 500 for that same cluster of central, eastern, and Balkan EU states. This is unexpected because, as noted, these regions experienced a wave October and November. Due to the steep age-risk gradient, NNIs declined markedly for individuals aged ≥60 such that even the median NNI was below 500 in most regions of the EU and UK. The NNIs were higher for critical illness (Figure 4) given that the ICRs were generally lower than the ISRs (Table S2). The Q1 of the NNIs for critical illness was ≥500 in the UK and most parts of the EU in all age-groups under 60. Lower NNIs were observed in the same cluster of central, eastern, and Balkan EU states.

**Figure 3.**
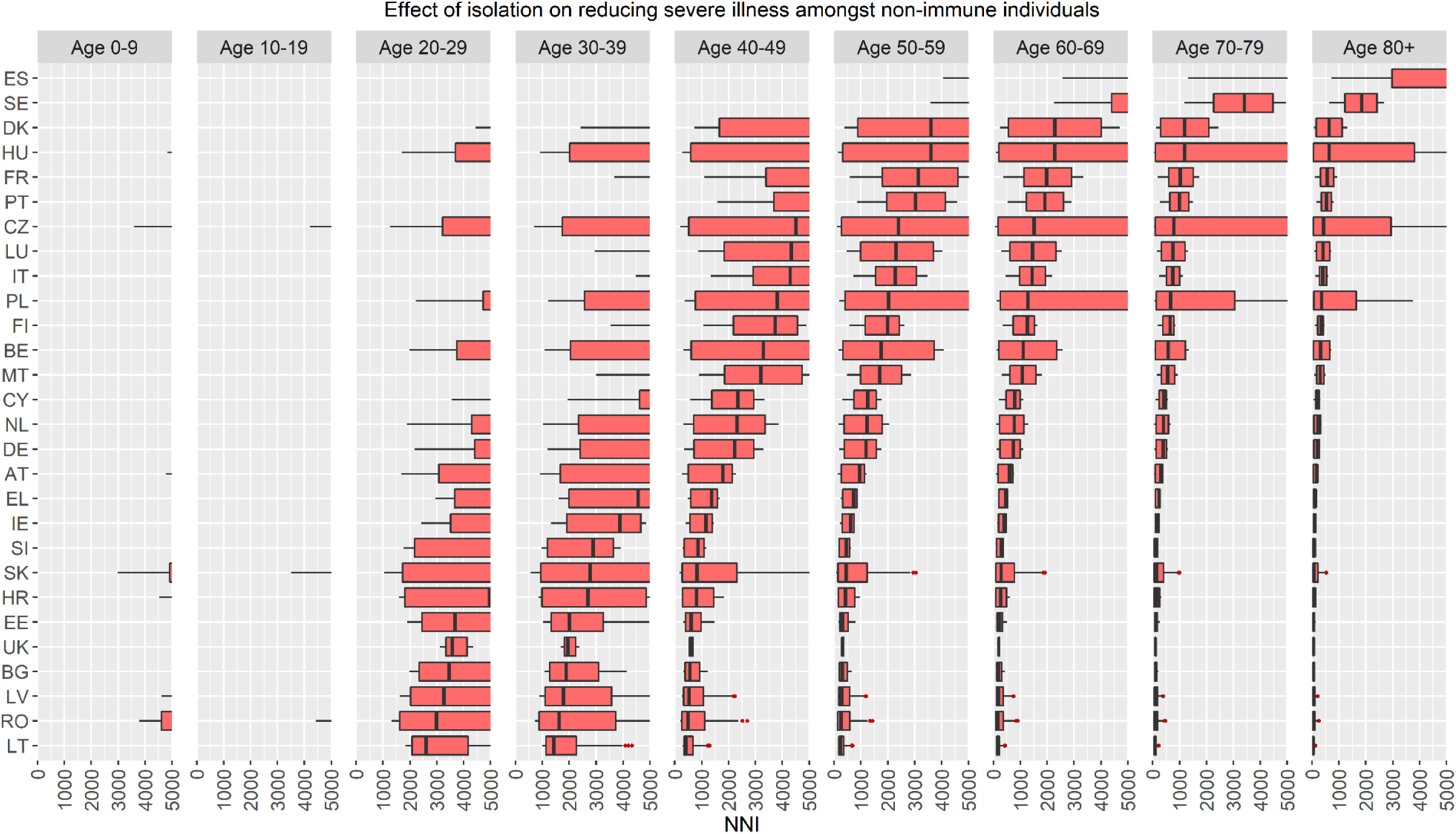
NNIs for severe illness in the European Union member states and United Kingdom. A region/age-group with no box-and-whisker plot has an NNI > 5,000 indicating a very low absolute risk reduction. AT=Austria, BE=Belgium, BG=Bulgaria, CY=Cyprus, CZ=Czechia, DE=Germany, DK=Denmark, EE=Estonia, EL=Greece, ES=Spain, FI=Finland, FR=France, HR=Croatia, HU=Hungary, IE=Ireland, IT=Italy, LT=Lithuania, LU=Luxembourg, LV=Latvia, MT=Malta, NL=Netherlands, PL=Poland, PT=Portugal, RO=Romania, SE=Sweden, SI=Slovenia, SK=Slovakia, UK=United Kingdom.

**Figure 4.**
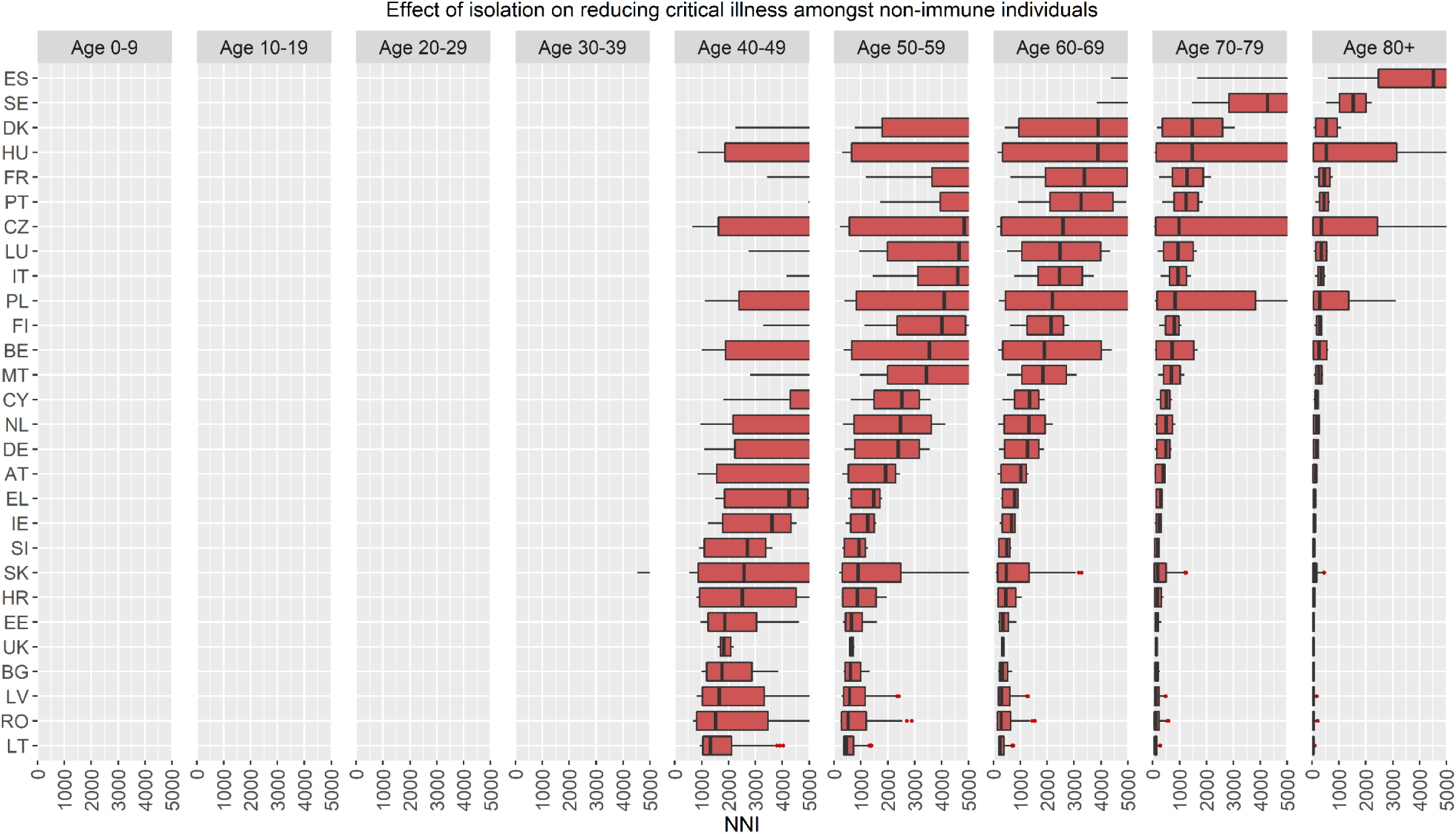
NNIs for critical illness in the European Union member states and United Kingdom. A region/age-group with no box-and-whisker plot has an NNI > 5,000 indicating a very low absolute risk reduction. AT=Austria, BE=Belgium, BG=Bulgaria, CY=Cyprus, CZ=Czechia, DE=Germany, DK=Denmark, EE=Estonia, EL=Greece, ES=Spain, FI=Finland, FR=France, HR=Croatia, HU=Hungary, IE=Ireland, IT=Italy, LT=Lithuania, LU=Luxembourg, LV=Latvia, MT=Malta, NL=Netherlands, PL=Poland, PT=Portugal, RO=Romania, SE=Sweden, SI=Slovenia, SK=Slovakia, UK=United Kingdom.

As described, the NNI is pushed higher or lower depending on if the IR is low or high, respectively. Therefore, while the pattern was similar in the other regions we examined, regions with IRs lower than the EU and UK had higher NNIs (e.g., Israel, Canada, Australia), whereas regions with IRs higher than the EU and UK had lower NNIs (e.g., many US states) (Figure S9).

In Israel, the Q1 of the NNI was ≥500 for transmission in all settings (Figure S10) and severe/critical illness in age-groups under 80 (Figures S11-S12). In Canada, the Q1 of the NNI was ≥500 for transmission in all settings and provinces/territories, except social gatherings in Saskatchewan, Northwest Territories, Alberta, and the Yukon (Figure S13), which are regions which suffered a wave in the fall 2021 (Figure S3). In Alberta, the Q1 for travel/transportation settings was also < 500. For severe illness, the Q1 was ≥500 in all age-groups under 60 in all parts of Canada except Saskatchewan, Northwest Territories, Alberta, and the Yukon (Figure S14). The same, but higher, pattern of NNIs for critical illness in Canada was found (Figure S15). In Australia, the Q1 of the NNI was ≥500 for transmission in all settings and states/territories, except social gatherings in Victoria (Figure S16) which experienced a small increase in IRs (∼2%) from October into November (Figure S4). The Q1 was ≥500 for severe/critical illness in almost all parts of Australia and age-groups (Figures S17-S18). For severe/critical illness, the exception was Victoria (ages ≥60-70) and New South Wales (ages ≥80). Note, many of the NNIs for transmission and severe/critical illness in Israel, Canada, and Australia were above 5,000, indicating very low ARRs of isolation on these outcomes on any given day during this period.

In the US Northeast, the Q1 of the NNIs for transmission were ≥500 for social gatherings only in Massachusetts, Connecticut, and Rhode Island. The Q1 was ≥500 in all Northeastern states for all other settings except New Hampshire for travel/transportation (Figure S19). Except for New Hampshire, it was ≥500 in ages under 50 for severe illness (Figure S20) and ages under 60 for critical illness (Figure S21). In the US Midwest, the Q1 for transmission in social gatherings was < 500 in all states (Figure S22). The Q1 was ≥500 for casual close contacts, healthcare, work/study places in all states except Nebraska. For travel/transportation settings, the Q1 was ≥500 in Iowa, Wisconsin, Indiana, South Dakota, Missouri, and Illinois. The Q1 for severe illness was ≥500 in all age-groups under 40 except Nebraska (Figure S23). A similar pattern was seen for critical illness (Figure S24). In the US South, the Q1 was ≥500 for transmission in social gatherings only in Maryland and Florida (Figure S25). The Q1 was ≥500 in all Southern states in casual close contacts, healthcare, work/study, and travel/transportation settings, except West Virginia, Kentucky, and Texas for travel/transportation. The Q1 was ≥500 in many Southern states in age-groups under 50 for severe illness and in age-groups under 60 for critical illness (Figures S26-27). In the US West, the Q1 for transmission in social gatherings was ≥500 only in Utah (Figure S28). For casual close contacts, healthcare, and work/study places, the Q1 was ≥500 except in Montana, Alaska, and Nevada. For travel/transportation, it was ≥500 only for Arizona, Oregon, Washington, California, Hawaii, and Utah. The Q1 was ≥500 in most Western states in ages under 40 for severe illness and ages under 50 for critical illness (Figures S29-S30).

## Discussion

This study found that the ARs of a non-household transmission event and a case of severe/critical illness in non-immune individuals were typically low on any given day in the fall 2021 in many of the countries we examined. As a result, the ARRs of isolation from September to November 2021 were low, leading to the high NNIs we observed (≥500). These NNIs indicate that, on any given day, one would have needed to isolate hundreds, and in some regions/age-groups thousands, of non-immune individuals from various settings to prevent one non-household transmission event or one case of severe/critical illness. For transmission, this was especially the case for casual close contact settings (e.g., public buildings/areas), healthcare settings, work/study places, and travel/transportation settings. For severe/critical illness, the ARRs were very small in individuals under 40 across all regions, resulting in very high NNIs (> 5,000). The NNI is not a fixed metric but rather time-varying based on the IR on a given day.

These high NNIs stem from the fact that the fall 2021 was a period when IRs were low (< 2%) and stable, with some exceptions (Figures S1-S8). These exceptions were regions which experienced waves (IRs=5-10%) (e.g., parts of the EU) or had sustained higher IRs (2-5%) (e.g., UK, US Northeast) during this period. In a context of higher IRs (≥2%), the ARRs of isolation on transmission from non-immune individuals in social gathering settings (e.g., gatherings of friends/family) was modest, such that the NNI was < 500. This was likewise for severe/critical illness amongst non-immune individuals aged ≥50-60. This latter finding is consistent with the steep age-risk gradient for SARS-CoV-2 and suggestions for measures focusing on protecting specific higher-risk age-groups [2]. Interestingly, even when IRs were sustained between 2-5%, the NNIs for the other non-household settings were ≥500.

These NNIs situate the ARRs of isolation using IMP within the range of the NNTs of ASA in primary prevention of CVD. This is salient since ASA is not recommended for primary prevention of CVD because the costs outweigh the benefits (NNTs ≥250) [16]. While not every region we examined implemented IMP during the fall 2021, some did with differing degrees of intensity (e.g., UK, EU states, Canada, Australia). Our findings suggest that, during a wave or sustained higher IRs, the ARRs of isolating non-immune individuals were modest for transmission only in intimate social gatherings and severe/critical illness in adults aged ≥50-60. When waves subsided and IRs fell below 2%, the ARRs were low and sometimes very low for non-household transmission and severe/critical illness. It is important to point out that NNIs ≥500 for non-household settings are for preventing one transmission event of an infection which is usually mild or asymptomatic. While the ISRs and ICRs are high (> 10%) for non-immune individuals aged ≥60 (Table S2), many of the countries we examined already had high levels of immunity from vaccination or prior infection by the time IMP were being discussed/implemented during this period. For example, the vaccination rate in these higher risk age-groups was 80-100% in the fall 2021 in many EU states [19]. This means the ISRs and ICRs in these high risk age-groups was likely significantly reduced by that time, given that the relative risk reductions of vaccination and prior infection for severe/critical illness are 80-95% and these benefits are robust over time [20-22].

The comparison with the NNTs of ASA in primary prevention of CVD is imperfect for two reasons. First, the NNT is primarily concerned with within-individual outcomes (e.g., myocardial infarction), whereas the NNI for non-household transmission concerns a between-individual outcome where one or more other individuals may be impacted (i.e., transmission event). Second, the ARR in the denominator of the NNI is based on point-prevalence, such that the risk is circumscribed over one day. The ARR in the denominator of the NNTs of ASA is often based on incidence proportions and pertains to risks over months and years. That said, we believe the comparison with the NNTs of ASA is helpful. This is because ASA is a good example in medicine of a clinical intuition that if one has to apply an intervention to hundreds or thousands of people to extract one benefit, a careful weighing of benefits vs. costs is needed.

A limitation of this analysis is that we had to extrapolate from wild-type data to estimate the ARs for Delta infections given that there was insufficient direct data. There is also likely a degree of underestimation in the DRDC database of the IRs. It is difficult to accurately model underreporting rates and how they change over time because one is trying to model something where there are no data. This is shown by the extreme variation in underreporting estimates [4, 23]. Local context/knowledge is required to estimate underreporting rates in a region over time, which is not available on a global scale. Estimating the NNIs required using publicly available data in published and preprint reports, such that several included studies have not yet been fully peer-reviewed. This was unavoidable due to the newly emerging evidence base on this topic and the multiple month lag from the peer-review process.

It is reasonable to ask why we did not use a risk metric to estimate the IR which uses a longer period of time (e.g., incidence proportion, period prevalence) since longer time windows would increase the IRs and thus lower the NNIs. Point-prevalence is the more appropriate metric for IR than incidence proportion and period prevalence for four reasons. First, the infection risk depends not just on new cases, but existing ones too. Second, incidence proportion and period prevalence depend on the time at risk. In general, shorter time windows will lower these metrics than longer time windows. If a time window is long enough, a cumulative risk can be high even if the risk on each day is low. However, there is no non-arbitrary way to set the time window to define the ‘correct’ time at risk. Time at risk is not an issue for point-prevalence because it is always a cross-section in time (the risk on a given day). Third, while risk over time is important, public health officials and communities are primarily concerned about the *current* risk of infection (i.e., point-prevalence), not, for example, the risk over the past 3 months. This was a retrospective study of September to November 2021, such that one could have taken the period prevalence of infectious cases during this time window to define the IRs. While this is not statistically incorrect to do, defining time at risk remains an issue. More importantly, period prevalence does not capture the risks that were known in the fall 2021 since, by definition, period prevalence is a retrospective measure (i.e., it is only known after the fact). Point-prevalence is known prospectively because it is a risk which can be measured on any given day.

Fourth, one could counter by pointing out that one can use forecasted risks to prospectively estimate IRs over longer time windows than one day. However, forecasted risks are not clearly preferable to point-prevalence. Inaccuracy and uncertainty of forecasts is a major challenge for any predictive model of infectious disease dynamics. This is because of the multifactorial/interacting nature of these dynamics and uncertainty in selecting/estimating the relevant predictors. Moreover, one cannot prospectively know if a forecasted IR is accurate since this, by definition, is discovered only retrospectively, which defeats the purpose of using forecasts to estimate NNIs. Relatedly, using period prevalence over a retrospective time window to forecast what the IR will be over the next months or years is challenging since it assumes the future will correspond to the past. The pandemic has shown that this is a tenuous assumption except over short periods of time. For these reasons, forecasted IRs are often more a form of speculation with wide uncertainty intervals than actually measurable risks.

Acknowledging these nuances and their impact on how to interpret the NNI, point-prevalence is the more appropriate metric of IR to estimate NNIs. The advantage is that point-prevalence is an actually measurable risk which can be known prospectively and does not suffer from time at risk issues. The disadvantage is that it does not quantify future risks or risks over longer periods of time. However, our data suggest that, on any given day during the fall 2021, the risks of a non-household transmission event or a severe/critical illness from non-immune individuals were typically low and sometimes very low. This means the NNIs were high, such that a careful weighing of benefits vs. costs was likely warranted.

## Supporting information

Supplementary Material - The NNI - Fall 2021

## Data Availability

All data are available in the manuscript/supplementary material and at https://github.com/TheNNIforViralTransmission/SARS-CoV-2 and https://covid-app.cloud.forces.gc.ca/map.

https://covid-app.cloud.forces.gc.ca/map

## Ethical approval

Not applicable.

## Contributions

AP, BH, and DS contributed equally to this work. AP, BH, and DS conceived the idea. AP and BH acquired the data, screened records, and extracted data. AP and BH performed the formal analysis. AP, BH, and DS wrote the first draft of the manuscript. All authors gave critical feedback on the revised report and approved the final version of the manuscript. The corresponding author attests all listed authors meet authorship criteria and that no others meeting criteria have been omitted.

## Acknowledgements

We thank numerous colleagues who provided helpful comments and criticisms during the writing of this study. We would especially like to thank Dr. Nathan Bakker and Dr. Mahesh Shenai for their valuable feedback on the ideas in this manuscript. This study received no grant from any funding agency, commercial, or not-for-profit sectors. It has also received no support of any kind from any individual or organization. BH is supported by a personal research grant from the University of Wroclaw within the “Excellence Initiative – Research University” framework and by a scholarship from the Polish Ministry of Education and Science. None of these institutions were involved in this research and did not fund it directly.

