## Supplementary Material - The NNI - Fall 2021 for "Estimating the risk reduction of isolation on COVID-19 non-household transmission and severe/critical illness in non-immune individuals: September to November 2021"

**Figure S26.** US South: NNIs for severe illness. 31
**Figure S27.** US South: NNIs for critical illness. 32
**Figure S28.** US West: NNIs for transmission in non-household settings. 33
**Figure S29.** US West: NNIs for severe illness. 34
**Figure S30.** US West: NNIs for critical illness. 35

**Appendix.** Search strategy and selection criteria to identify meta-analyses of the wild-type secondary attack rates (SARs) for SARS-CoV-2.

To identify the meta-analyses used to estimate the wild-type SARs, PubMed was searched from inception to January 9th, 2022, with no language restrictions. We used the search terms "SARS-CoV-2" or "COVID-19" combined with “secondary attack*” and “meta-analysis”. Two authors (AP, BH) screened the titles/abstracts to determine potential eligibility. Full-texts of potentially eligible reports were retrieved and assessed to determine eligibility. Data were independently extracted. Disagreements at either the screening, full-text review, or data extraction stages were resolved through consensus. Reports were included if they were a meta-analysis of studies of SARs from 2020 and estimated the SAR for at least one of six types of settings: (1) households, (2) social gatherings (e.g., meals, conversations, social gatherings with friends/family), (3) casual close contacts (e.g., public areas or buildings), (4) work or study places, (5) healthcare, and (6) travel/transportation. Meta-analyses of studies during the vaccine period (2021) were excluded to restrict the data to non-immune index cases. Random effect model data were extracted whenever possible. 15 citations were identified from the search. 1 meta-analysis [1] was known to our group. The meta-analytic estimates of the SARs were extracted from the 7 meta-analyses meeting the inclusion criteria [1-7]. The raw data of the wild-type SARs are available online [8]. All analyses were conducted in R [9].

**Table S1.** Estimates of the secondary attack rates used to estimate the absolute risk of a transmission event in non-household settings from a non-immune person infected with the Delta variant.

| **Type of setting** | **Wild-type SAR ($SAR_{wt}$)** | **Delta SAR ($SAR_{Delta}$): $SAR_{wt} \times CF_{Delta}$** |
|---|---|---|
| Households | 16.54% (13.73%, 19.36%) | 32.59% (27.04%, 38.14%) |
| Social gatherings | 5.93% (5.87%, 6.00%) | 11.69% (11.56%, 11.82%) |
| Casual close contacts | 1.55% (0.86%, 2.24%) | 3.05% (1.70%, 4.40%) |
| Work/study places | 1.47% (0.96%, 1.98%) | 2.89% (1.88%, 3.89%) |
| Healthcare | 1.50% (0.12%, 2.88%) | 2.96% (0.24%, 5.67%) |
| Travel/transportation | 2.23% (0.00%, 4.95%) | 4.40% (0.00%, 9.75%) |

Brackets are the 95% confidence intervals around the mean (lower limit, upper limit). SAR=secondary attack rate. Note, the lower limit of the SAR for travel/transportation crossed zero and thus it was set to zero. $CF_{Delta}$=1.97.

**Table S2.** Estimates of the severe and critical illness rates used to estimate the absolute risk of a severe/critical illness in a non-immune person infected with the Delta variant.

| **Age-group** | **Wild-type ISR ($ISR_{wt}$)** | $CF_{Delta}$ | **Delta ISR ($ISR_{Delta}$): $ISR_{wt} \times CF_{Delta}$** | **Wild-type ICR ($ICR_{wt}$)** | $CF_{Delta}$ | **Delta ICR ($ICR_{Delta}$): $ICR_{wt} \times CF_{Delta}$** |
|---|---|---|---|---|---|---|
| **0-9** | 0.103% (0.063%, 0.162%) | 2.51 | 0.259% (0.158%, 0.407%) | 0.0088% (0.0053%, 0.0139%) | 1 | 0.0088% (0.0053%, 0.0139%) |
| **10-19** | 0.22% (0.13%, 0.35%) | 1 | 0.22% (0.13%, 0.35%) | 0.024% (0.014%, 0.037%) | 1 | 0.024% (0.014%, 0.037%) |
| **20-29** | 0.47% (0.28%, 0.74%) | 1.57 | 0.74% (0.44%, 1.16%) | 0.063% (0.038%, 0.10%) | 1 | 0.063% (0.038%, 0.10%) |
| **30-39** | 0.99% (0.57%, 1.61%) | 1.37 | 1.36% (0.78%, 2.21%) | 0.17% (0.10%, 0.28%) | 1 | 0.17% (0.10%, 0.28%) |
| **40-49** | 2.1% (1.2%, 3.5%) | 2.16 | 4.5% (2.6%, 7.6%) | 0.46% (0.26%, 0.77%) | 3.17 | 1.46% (0.82%, 2.44%) |
| **50-59** | 4.4% (2.4%, 7.4%) | 1.94 | 8.5% (4.7%, 14.4%) | 1.2% (0.6%, 2.1%) | 3.52 | 4.2% (2.1%, 7.4%) |
| **60-69** | 8.9% (4.6%, 15.2%) | 1.52 | 13.5% (7.0%, 23.1%) | 3.3% (1.6%, 5.9%) | 2.4 | 7.9% (3.8%, 14.2%) |
| **70-79** | 17.1% (8.9%, 28.8%) | 1.53 | 26.2% (13.6%, 44.1%) | 8.3% (3.9%, 15.5%) | 2.52 | 20.9% (9.8%, 39.1%) |
| **≥ 80** | 30.3% (16.4%, 47.7%) | 1.6 | 48.5% (26.2%, 76.3%) | 19.4% (9.2%, 34.7%) | 3.01 | 58.4% (27.7%, 100%) |

Brackets are the 95% credibility intervals around the mean (lower limit, upper limit). ISR=infection-severe rate. ICR=infection-critical rate. The age-stratified wild-type ISRs and ICRs are from Herrera-Esposito & de los Campos (2021) who calculated credibility intervals around these estimates [10]. $CF_{Delta}$=correction factor for the increased severity of the Delta variant vs. wild-type using age-stratified adjusted odds ratios for hospitalisation (ISR) and ICU admission (ICR) [11]. If there was no statistically significant difference in the age-group, the value was set to 1.

**Figure S1.** Time trends in infection risks in the European Union member states and United Kingdom from September to November 2021.

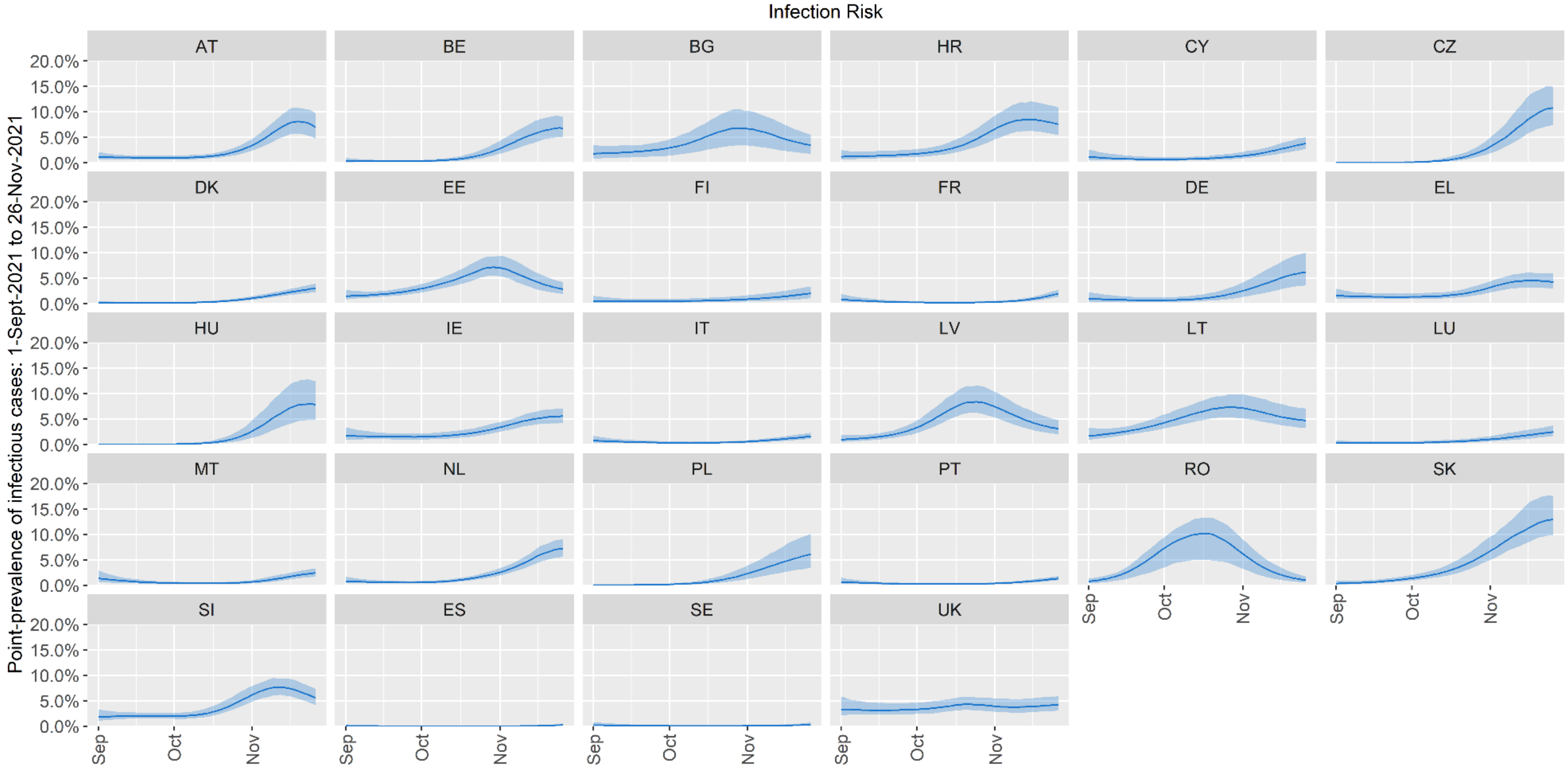

Error bars are 95% confidence intervals. AT=Austria, BE=Belgium, BG=Bulgaria, CY=Cyprus, CZ=Czechia, DE=Germany, DK=Denmark, EE=Estonia, EL=Greece, ES=Spain, FI=Finland, FR=France, HR=Croatia, HU=Hungary, IE=Ireland, IT=Italy, LT=Lithuania, LU=Luxembourg, LV=Latvia, MT=Malta, NL=Netherlands, PL=Poland, PT=Portugal, RO=Romania, SE=Sweden, SI=Slovenia, SK=Slovakia, UK=United Kingdom.

**Figure S2.** Time trends in infection risks in Israel from September to November 2021.

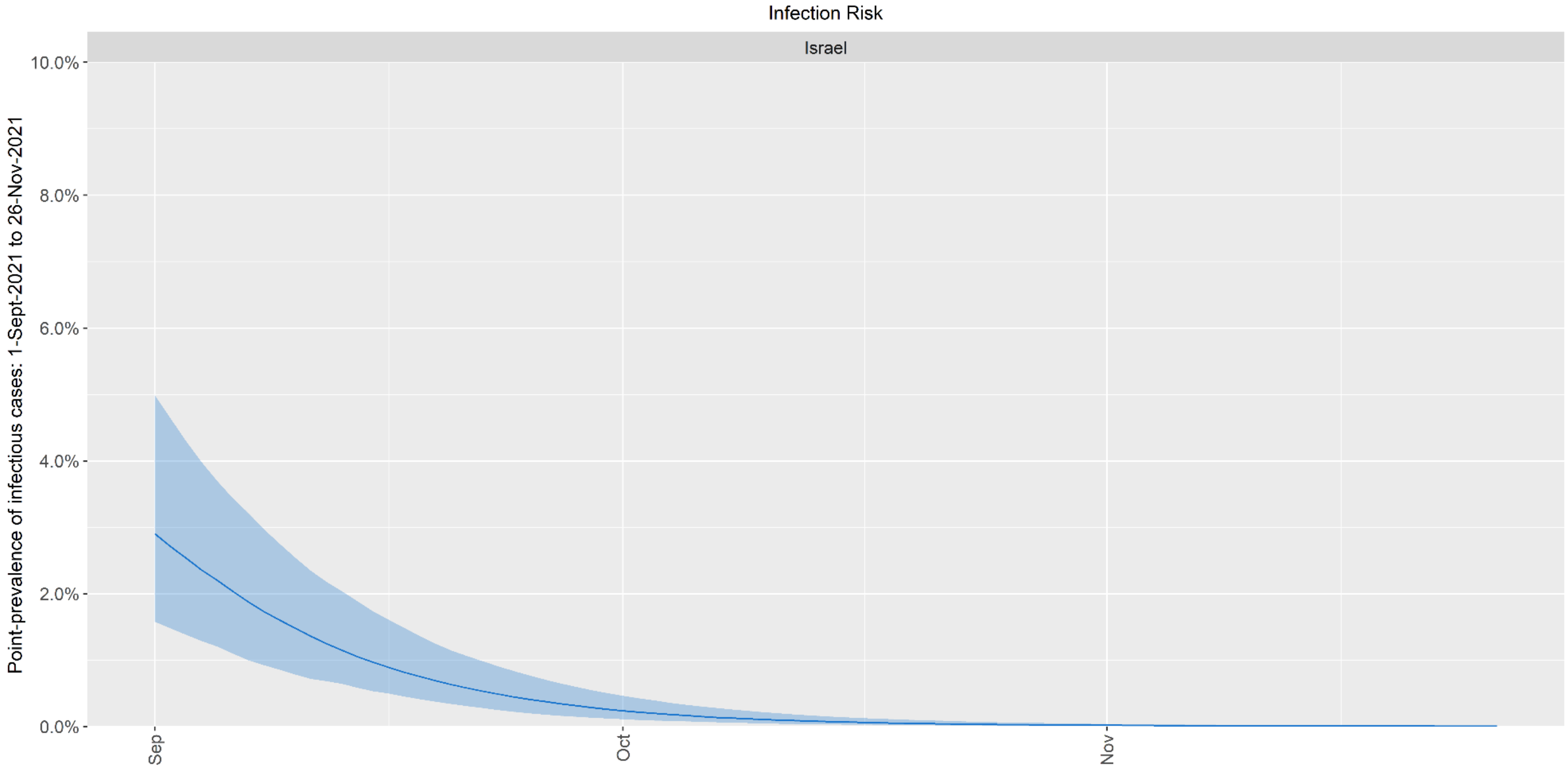

Error bars are 95% confidence intervals.

**Figure S3.** Time trends in infection risks in Canada from September to November 2021.

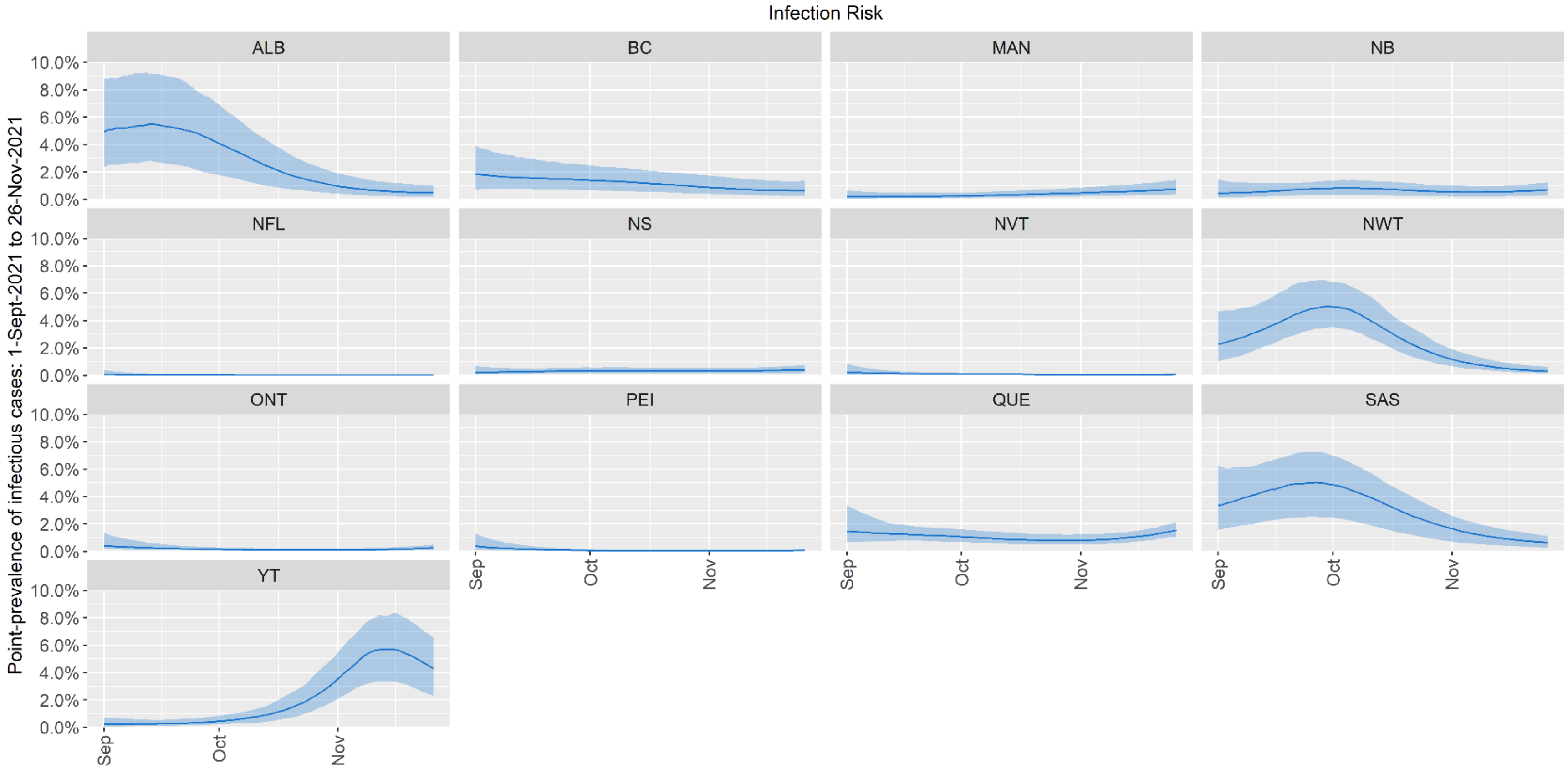

Error bars are 95% confidence intervals. ALB=Alberta, BC=British Columbia, MAN=Manitoba, NB=New Brunswick, NFL=Newfoundland and Labrador, NS=Nova Scotia, NVT=Nunavut, NWT=Northwest Territories, ONT=Ontario, PEI=Prince Edward Island, QUE=Quebec, SAS=Saskatchewan, YT=Yukon.

**Figure S4.** Time trends in infection risks in Australia from September to November 2021.

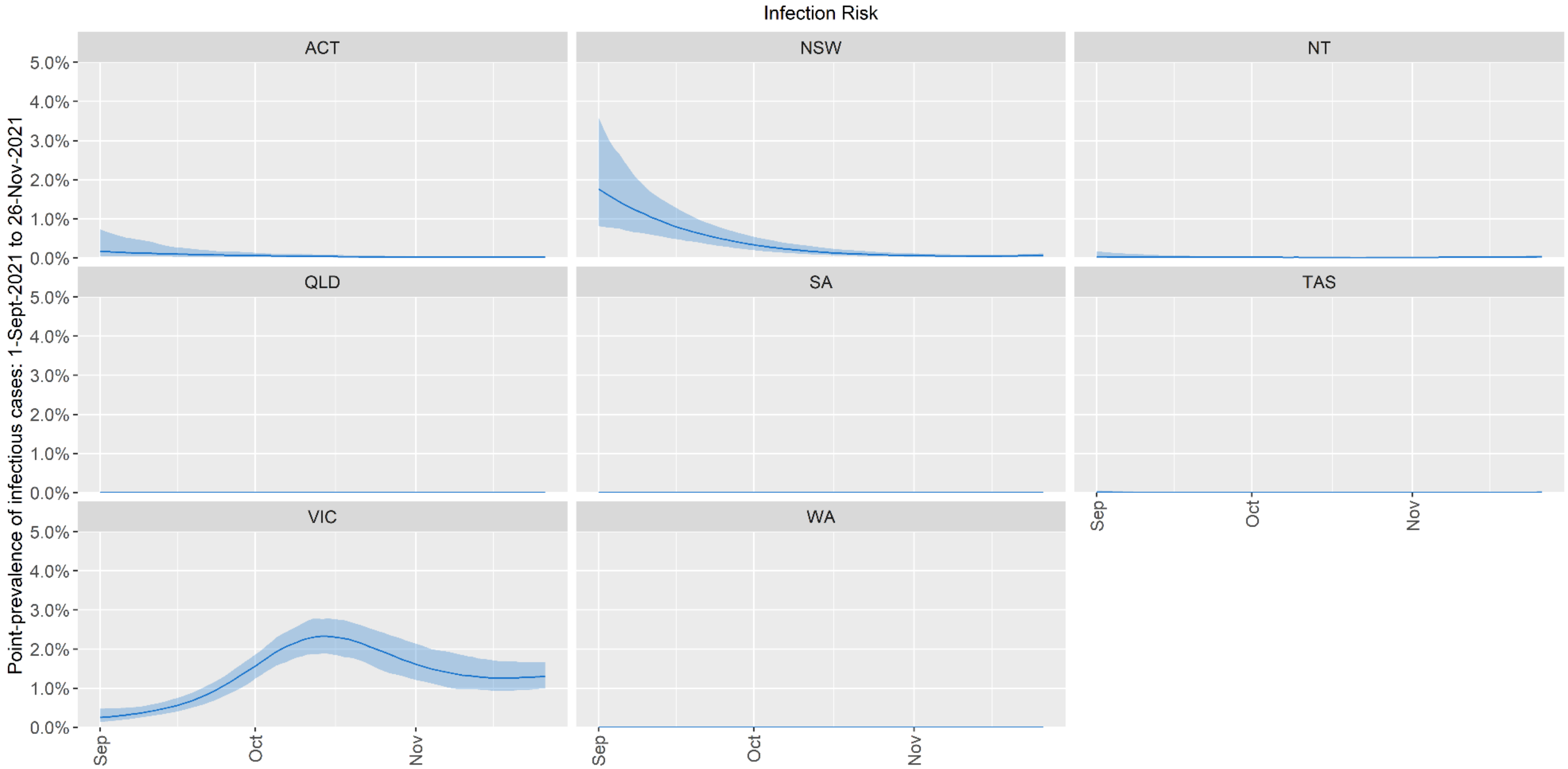

Error bars are 95% confidence intervals. ACT=Australian Capital Territory, NSW=New South Wales, NT=Northern Territory, QLD=Queensland, SA=South Australia, TAS=Tasmania, VIC=Victoria, WA=Western Australia.

**Figure S5.** Time trends in infection risks in the US Northeast from September to November 2021.

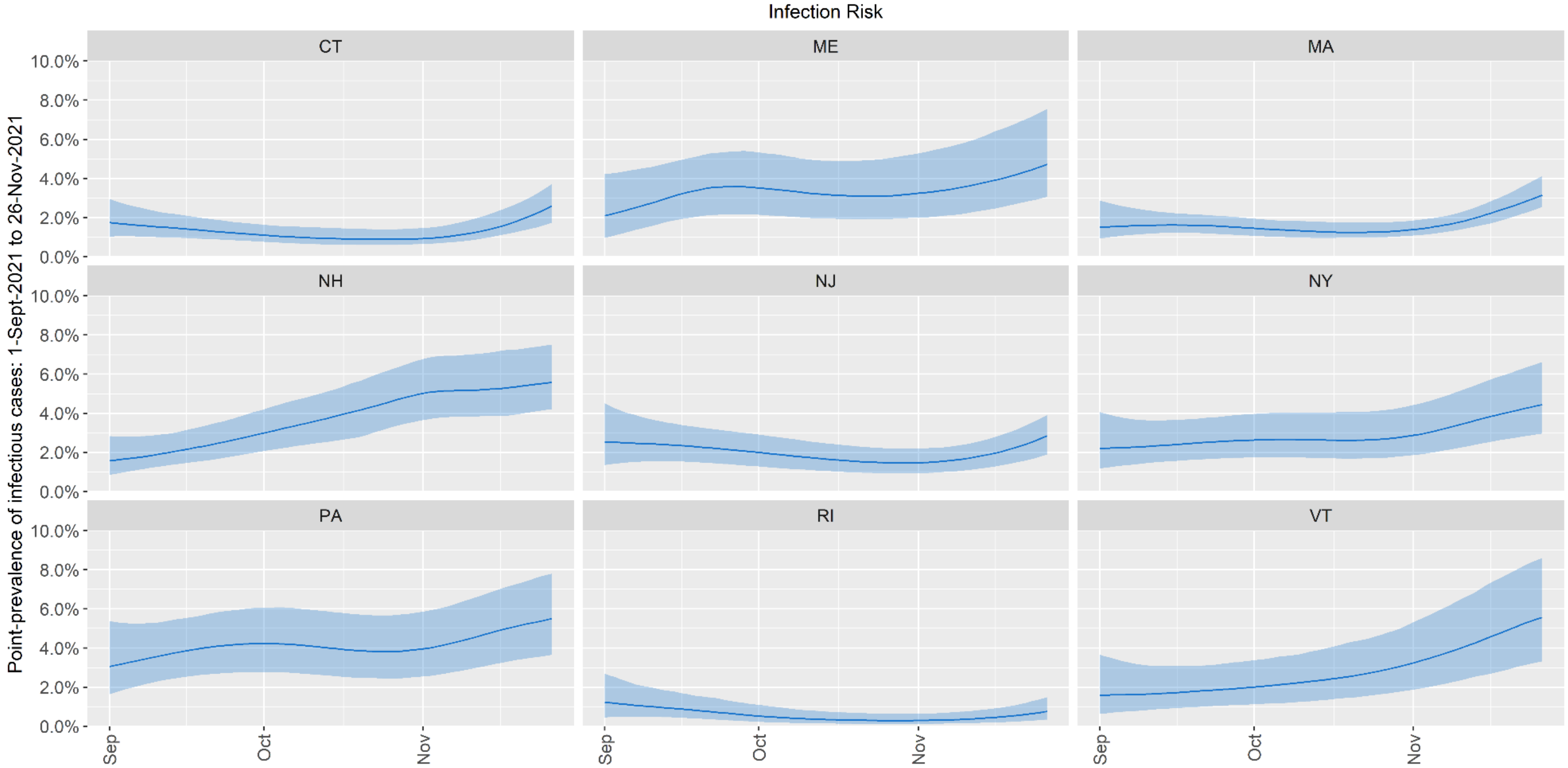

Error bars are 95% confidence intervals. CT=Connecticut, ME=Maine, MA=Massachusetts, NH=New Hampshire, NJ=New Jersey, NY=New York, PA=Pennsylvania, RI=Rhode Island, VT=Vermont.

**Figure S6.** Time trends in infection risks in the US Midwest from September to November 2021.

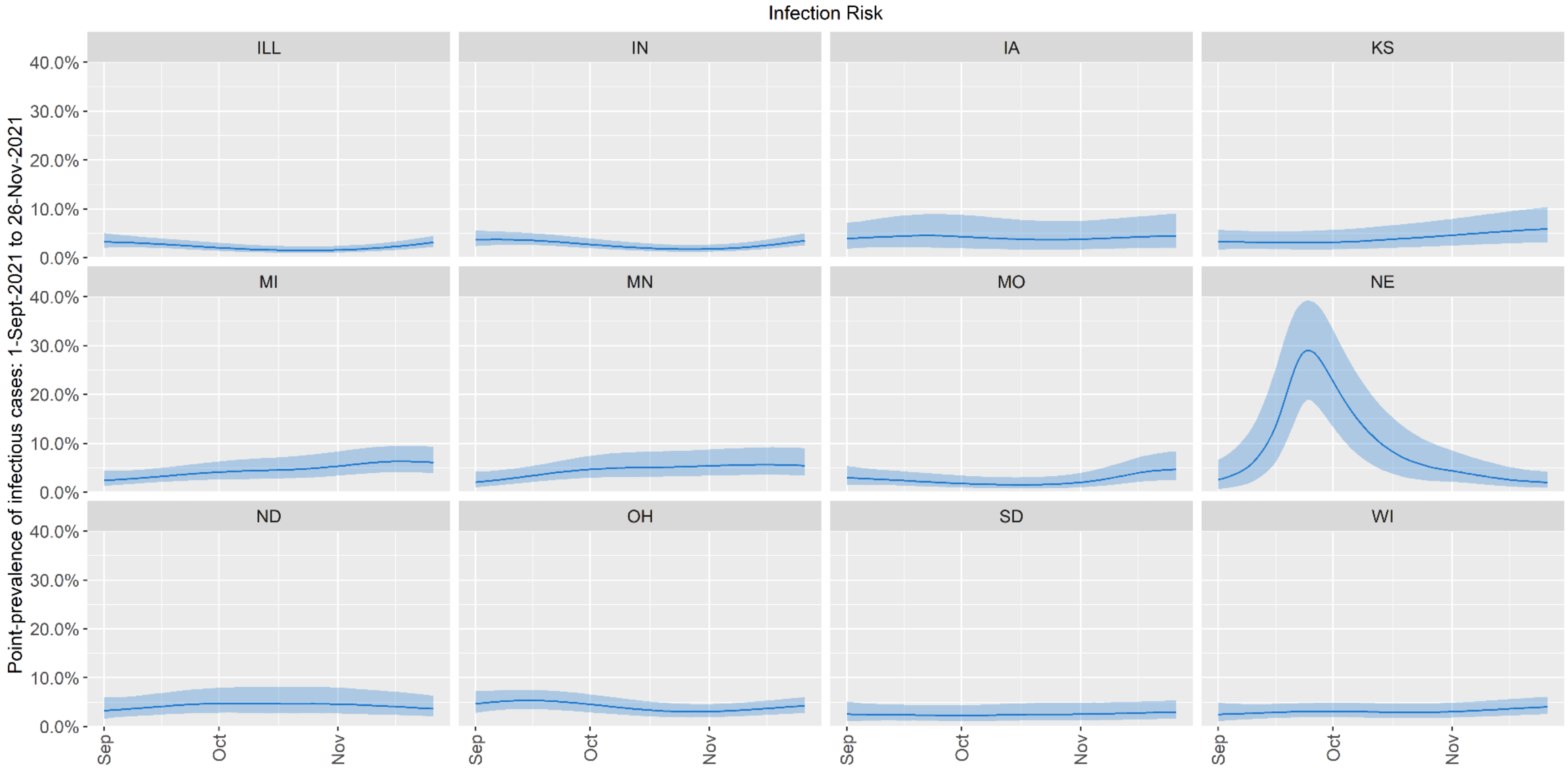

Error bars are 95% confidence intervals. ILL=Illinois, IN=Indiana, IA=Iowa, KS=Kansas, MI=Michigan, MN=Minnesota, MO=Missouri, NE=Nebraska, ND=North Dakota, OH=Ohio, SD=South Dakota, WI=Wisconsin.

**Figure S7.** Time trends in infection risks in the US South from September to November 2021.

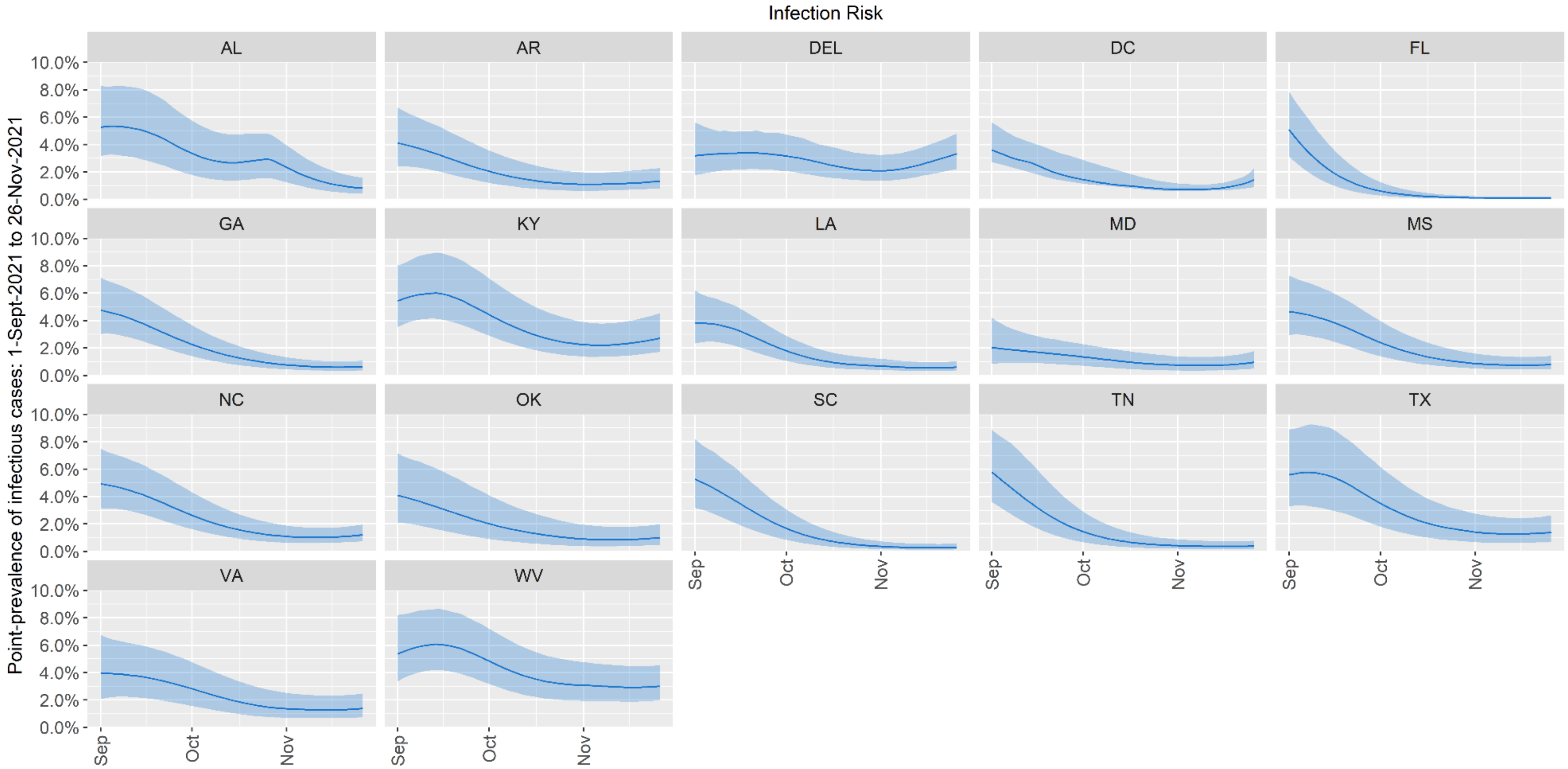

Error bars are 95% confidence intervals. AL=Alabama, AR=Arkansas, DEL=Delaware, DC=District of Columbia, FL=Florida, GA=Georgia, KY=Kentucky, LA=Louisiana, MD=Maryland, MS=Mississippi, NC=North Carolina, OK=Oklahoma, SC=South Carolina, TN=Tennessee, TX=Texas, VA=Virginia, WV=West Virginia.

**Figure S8.** Time trends in infection risks in the US West from September to November 2021.

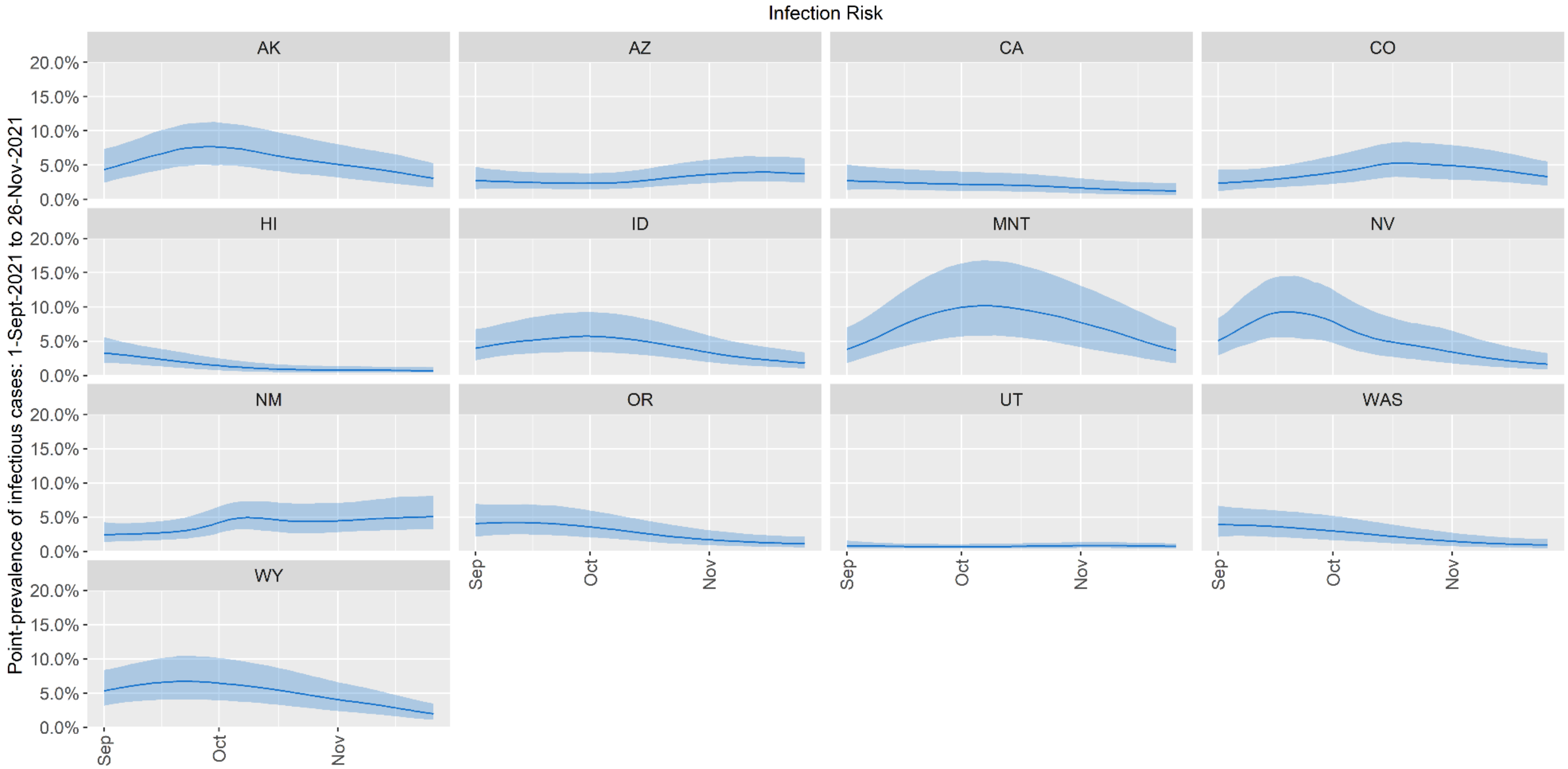

Error bars are 95% confidence intervals. AK=Alaska, AZ=Arizona, CA=California, CO=Colorado, HI=Hawaii, ID=Idaho, MNT=Montana, NV=Nevada, NM=New Mexico, OR=Oregon, UT=Utah, WAS=Washington, WY=Wyoming.

**Figure S9.** Daily infection risks from September to November 2021 by major jurisdiction.

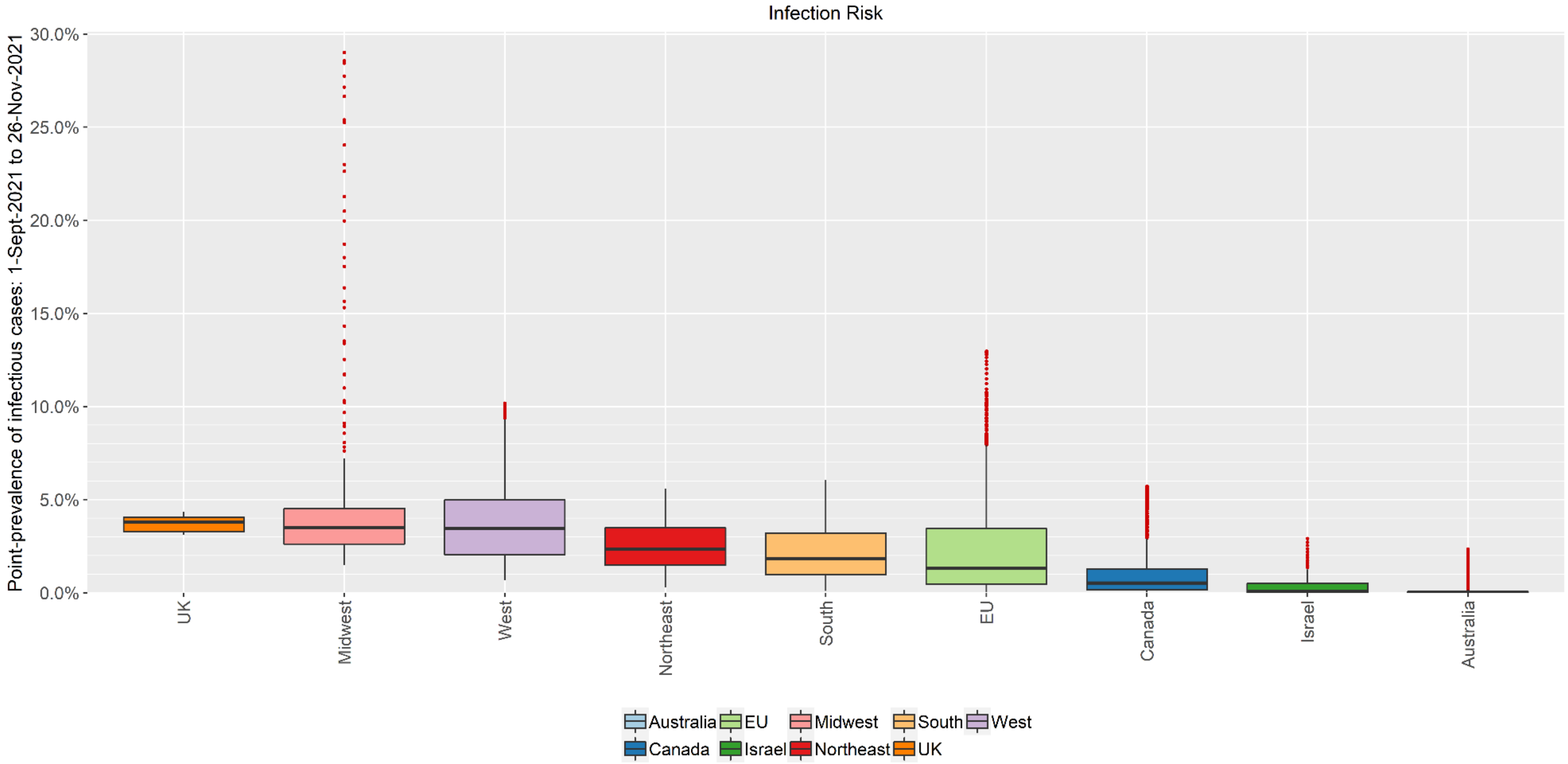

Major jurisdictions examined were EU member states, the United Kingdom (UK), Israel, Canada, Australia, and the major US regions (Northeast, Midwest, South, West). The distribution of risks during this period are displayed using box-and-whisker plots.

**Figure S10.** Israel: NNIs for transmission in non-household settings.

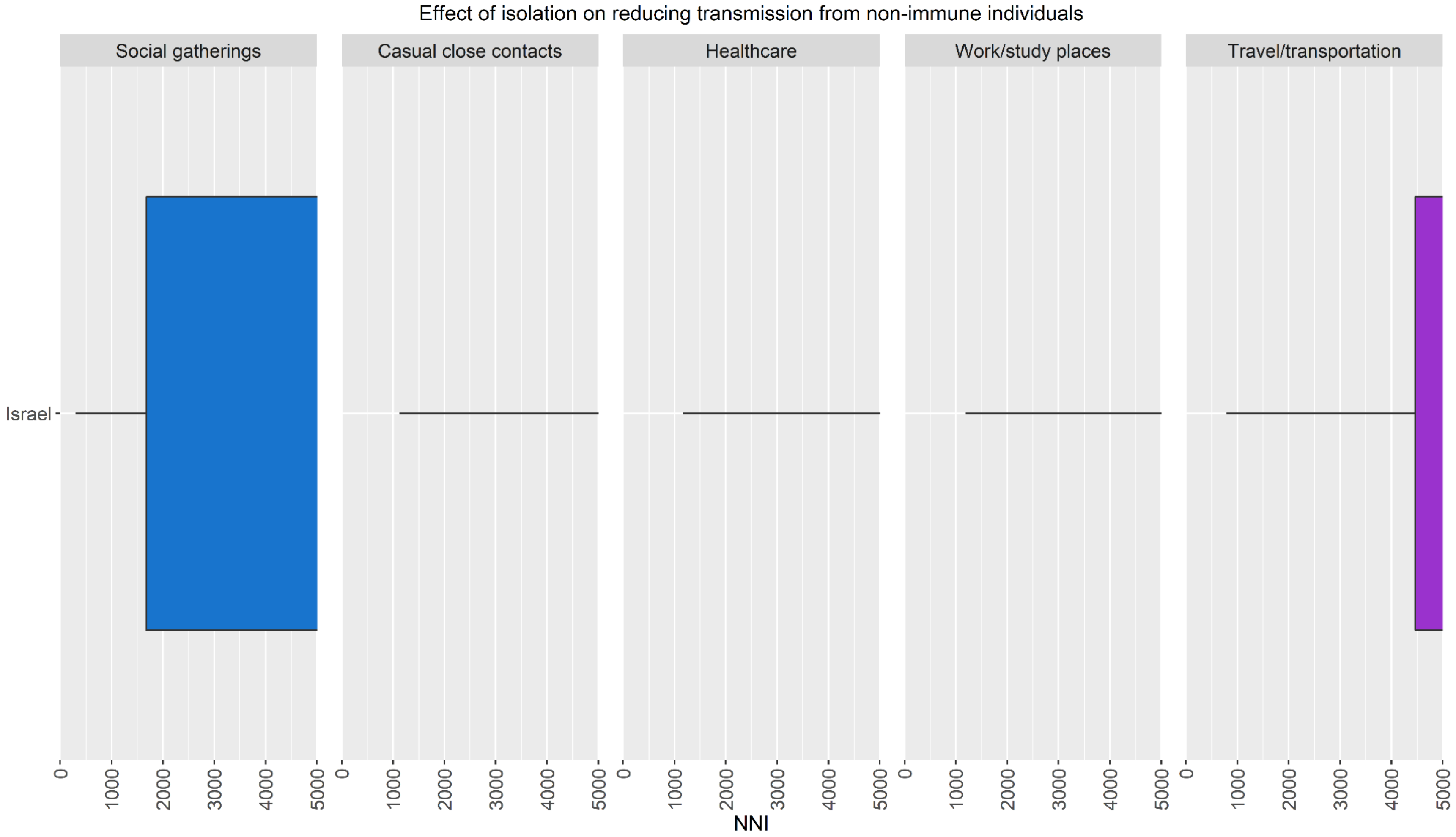

A setting with no box-and-whisker plot has an NNI > 5,000 indicating a very low absolute risk reduction.

**Figure S11.** Israel: NNIs for severe illness.

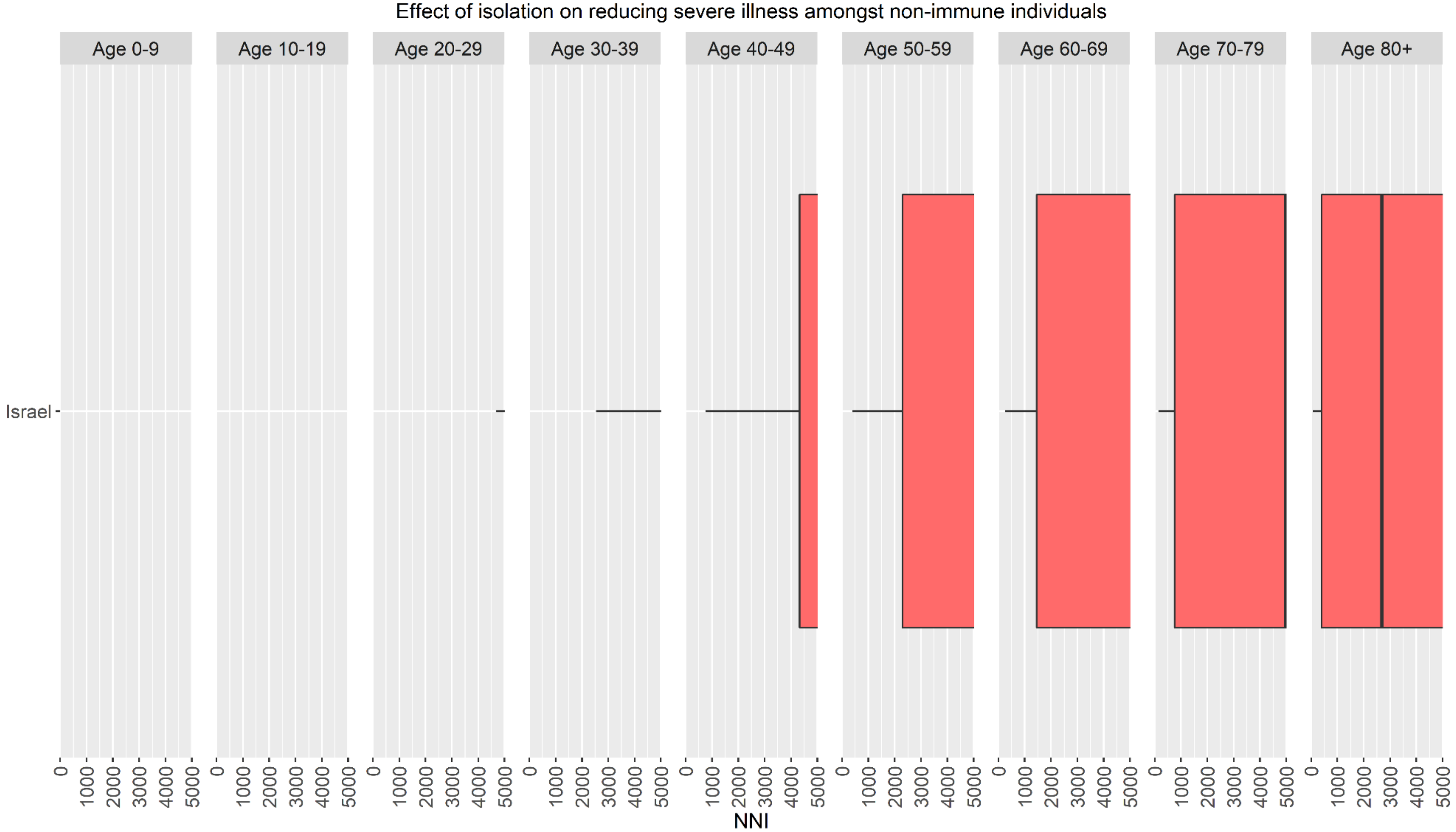

An age-group with no box-and-whisker plot has an NNI > 5,000 indicating a very low absolute risk reduction.

**Figure S12.** Israel: NNIs for critical illness.

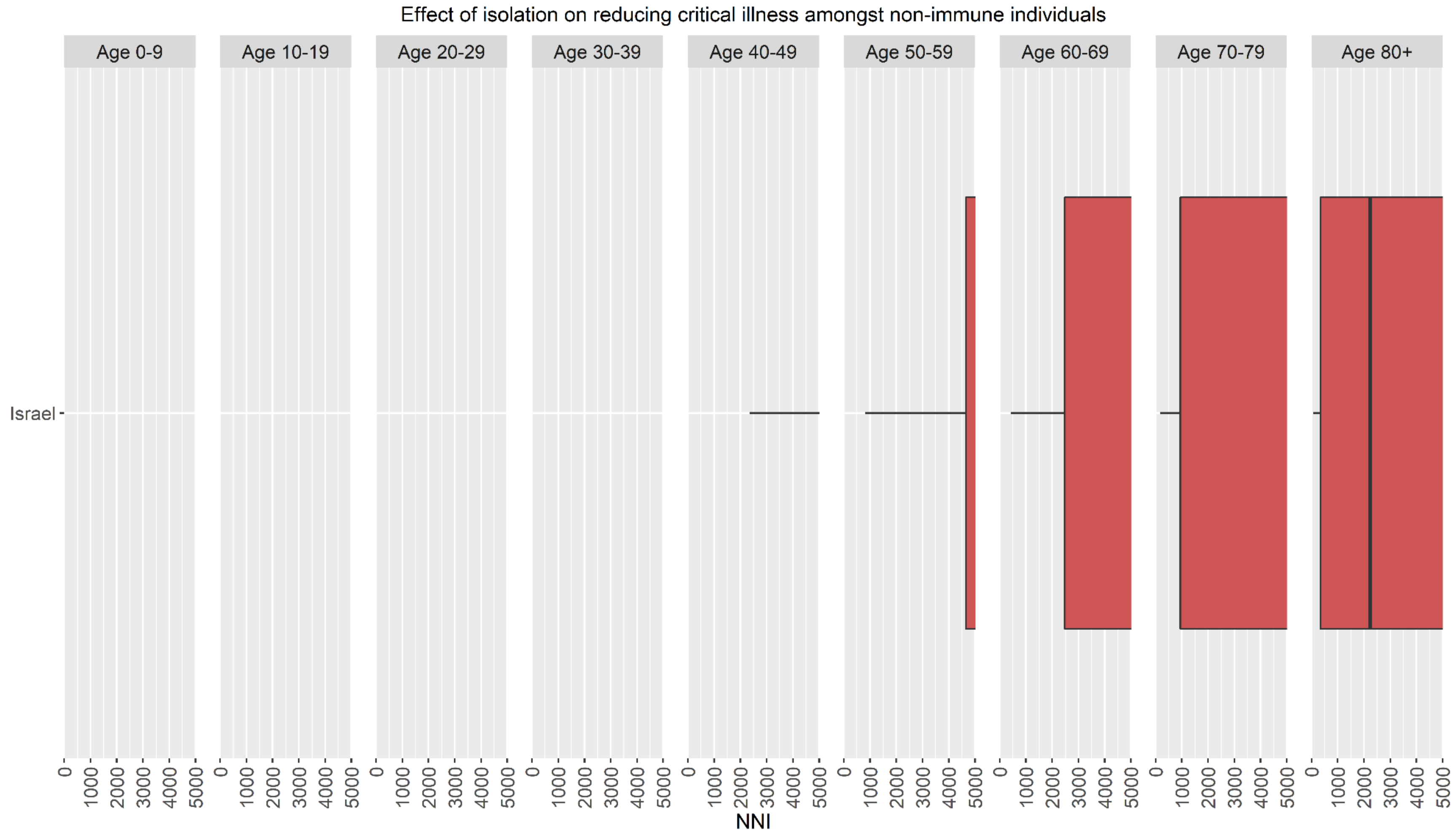

An age-group with no box-and-whisker plot has an NNI > 5,000 indicating a very low absolute risk reduction.

**Figure S13.** Canada: NNIs for transmission in non-household settings.

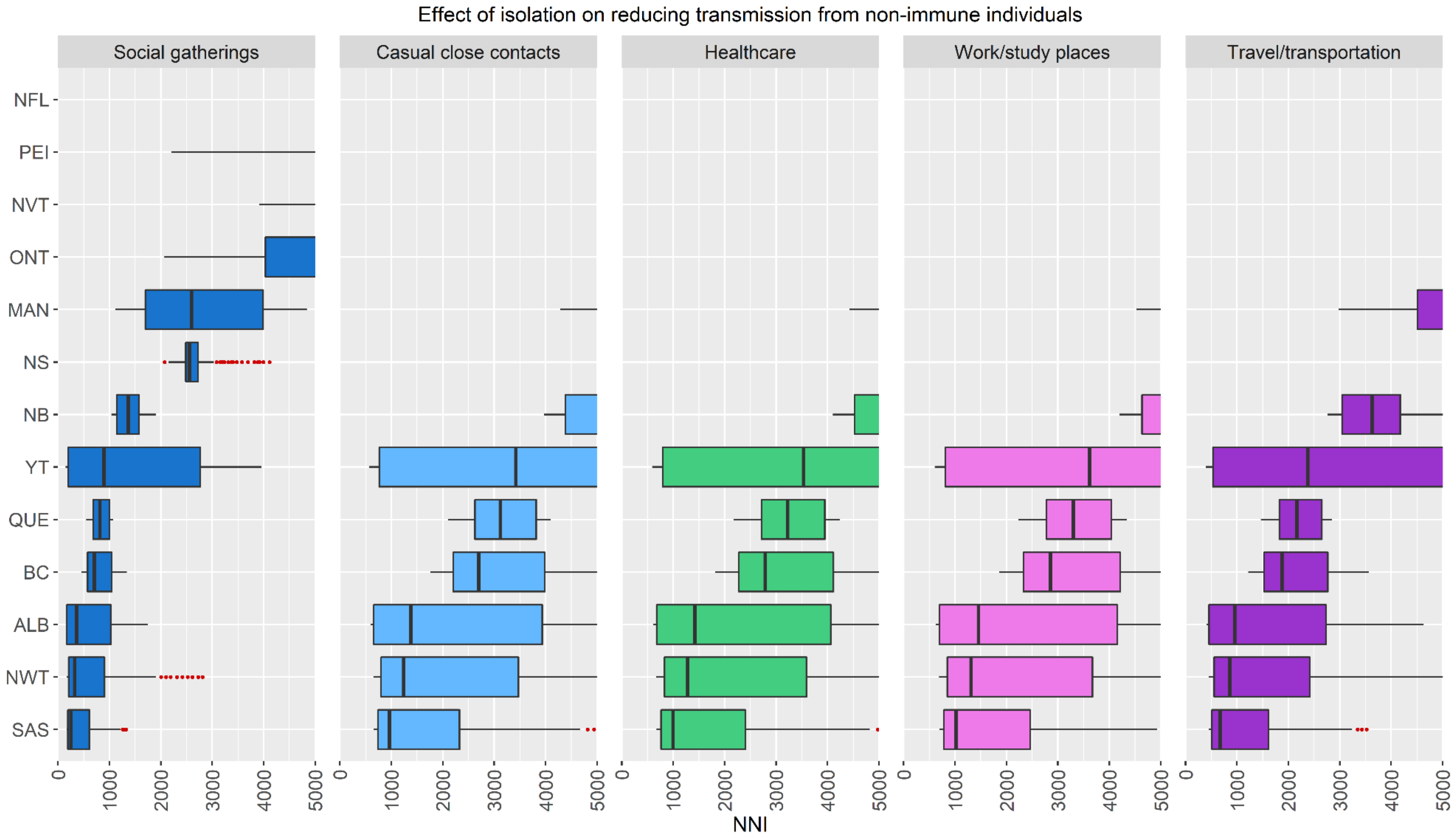

A region/setting with no box-and-whisker plot has an NNI > 5,000 indicating a very low absolute risk reduction. ALB=Alberta, BC=British Columbia, MAN=Manitoba, NB=New Brunswick, NFL=Newfoundland and Labrador, NS=Nova Scotia, NVT=Nunavut, NWT=Northwest Territories, ONT=Ontario, PEI=Prince Edward Island, QUE=Quebec, SAS=Saskatchewan, YT=Yukon.

**Figure S14.** Canada: NNIs for severe illness.

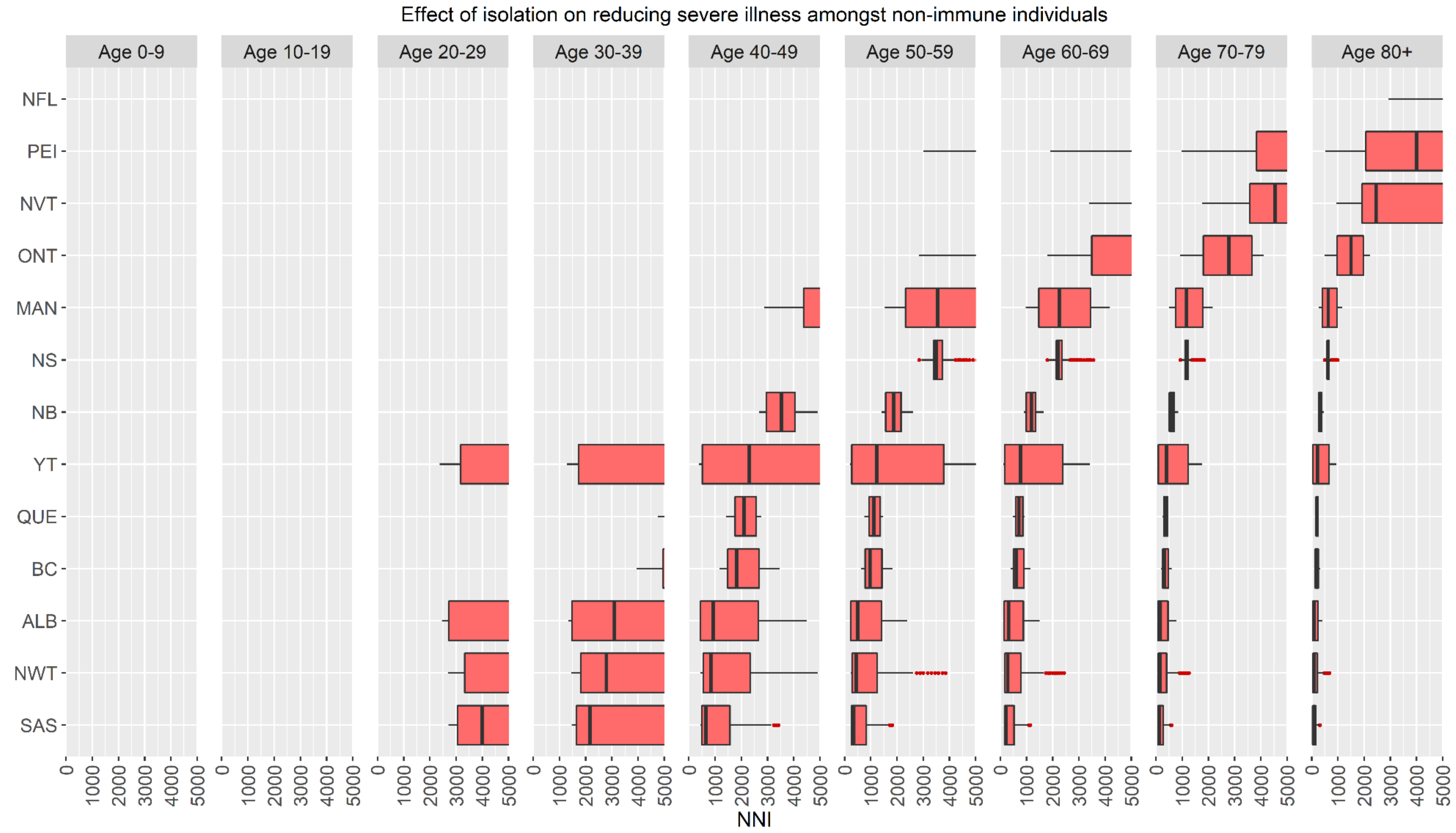

A region/age-group with no box-and-whisker plot has an NNI > 5,000 indicating a very low absolute risk reduction. ALB=Alberta, BC=British Columbia, MAN=Manitoba, NB=New Brunswick, NFL=Newfoundland and Labrador, NS=Nova Scotia, NVT=Nunavut, NWT=Northwest Territories, ONT=Ontario, PEI=Prince Edward Island, QUE=Quebec, SAS=Saskatchewan, YT=Yukon.

**Figure S15.** Canada: NNIs for critical illness.

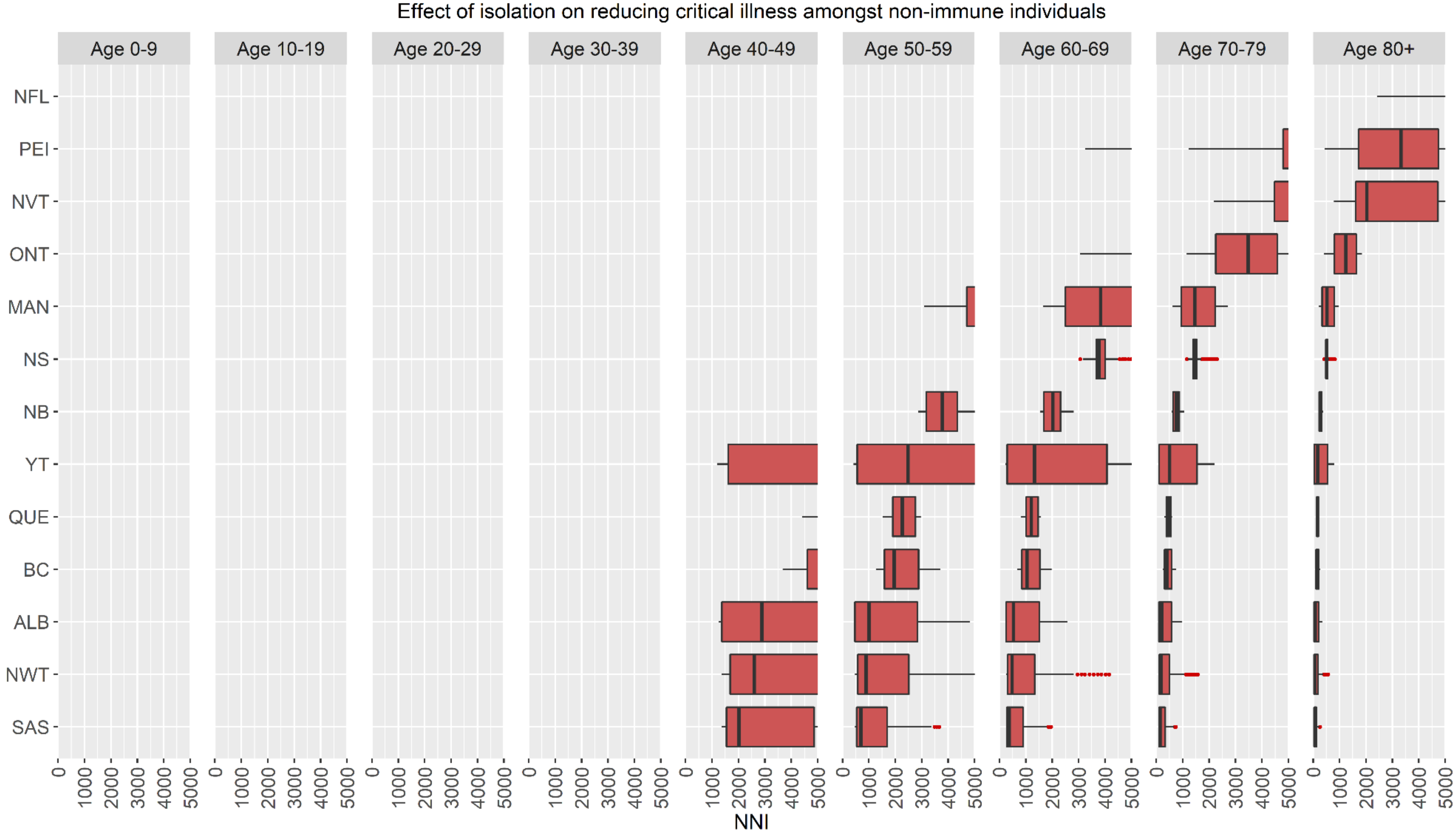

A region/age-group with no box-and-whisker plot has an NNI > 5,000 indicating a very low absolute risk reduction. ALB=Alberta, BC=British Columbia, MAN=Manitoba, NB=New Brunswick, NFL=Newfoundland and Labrador, NS=Nova Scotia, NVT=Nunavut, NWT=Northwest Territories, ONT=Ontario, PEI=Prince Edward Island, QUE=Quebec, SAS=Saskatchewan, YT=Yukon.

**Figure S16.** Australia: NNIs for transmission in non-household settings.

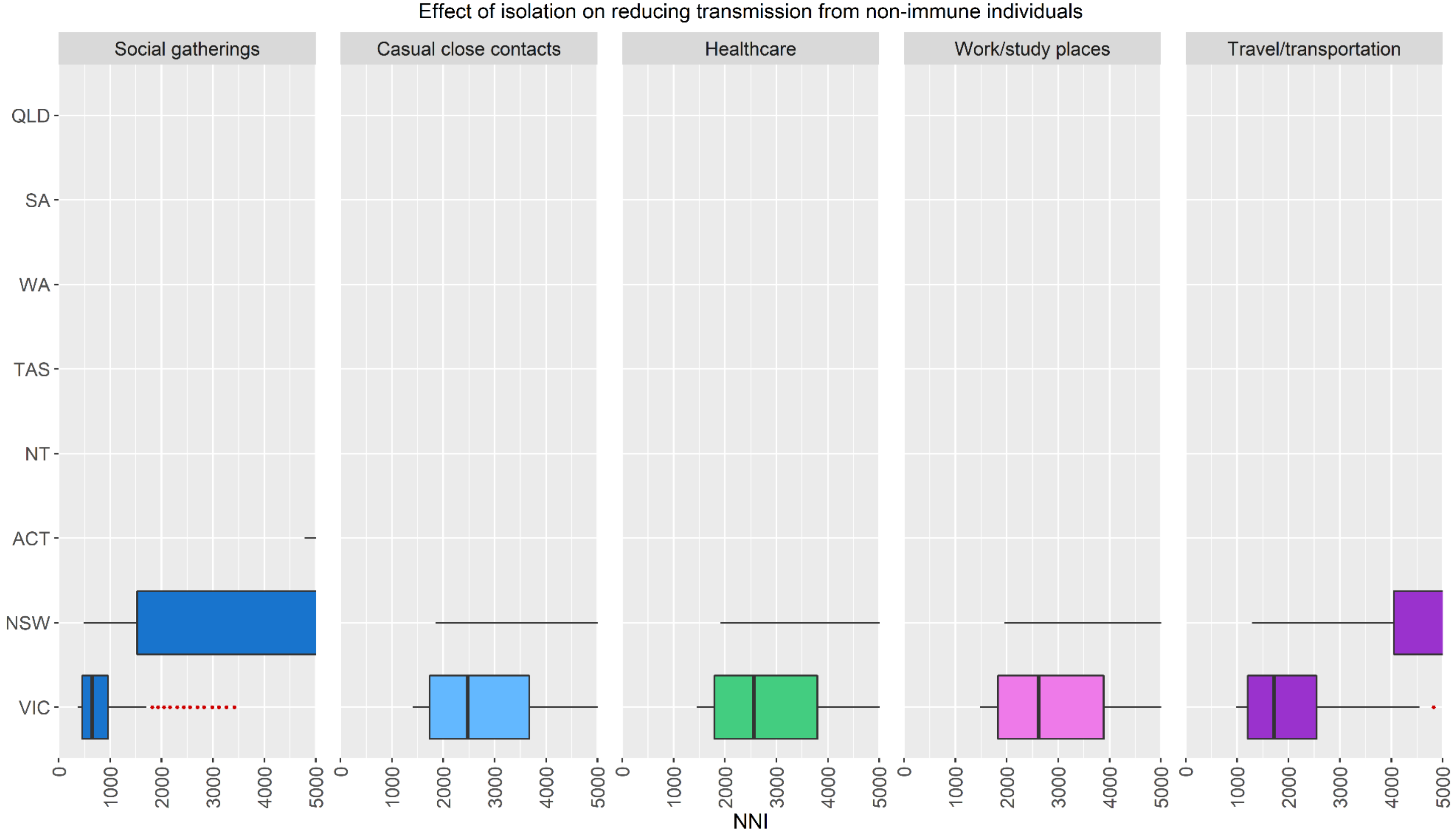

A region/setting with no box-and-whisker plot has an NNI > 5,000 indicating a very low absolute risk reduction. ACT=Australian Capital Territory, NSW=New South Wales, NT=Northern Territory, QLD=Queensland, SA=South Australia, TAS=Tasmania, VIC=Victoria, WA=Western Australia.

**Figure S17.** Australia: NNIs for severe illness.

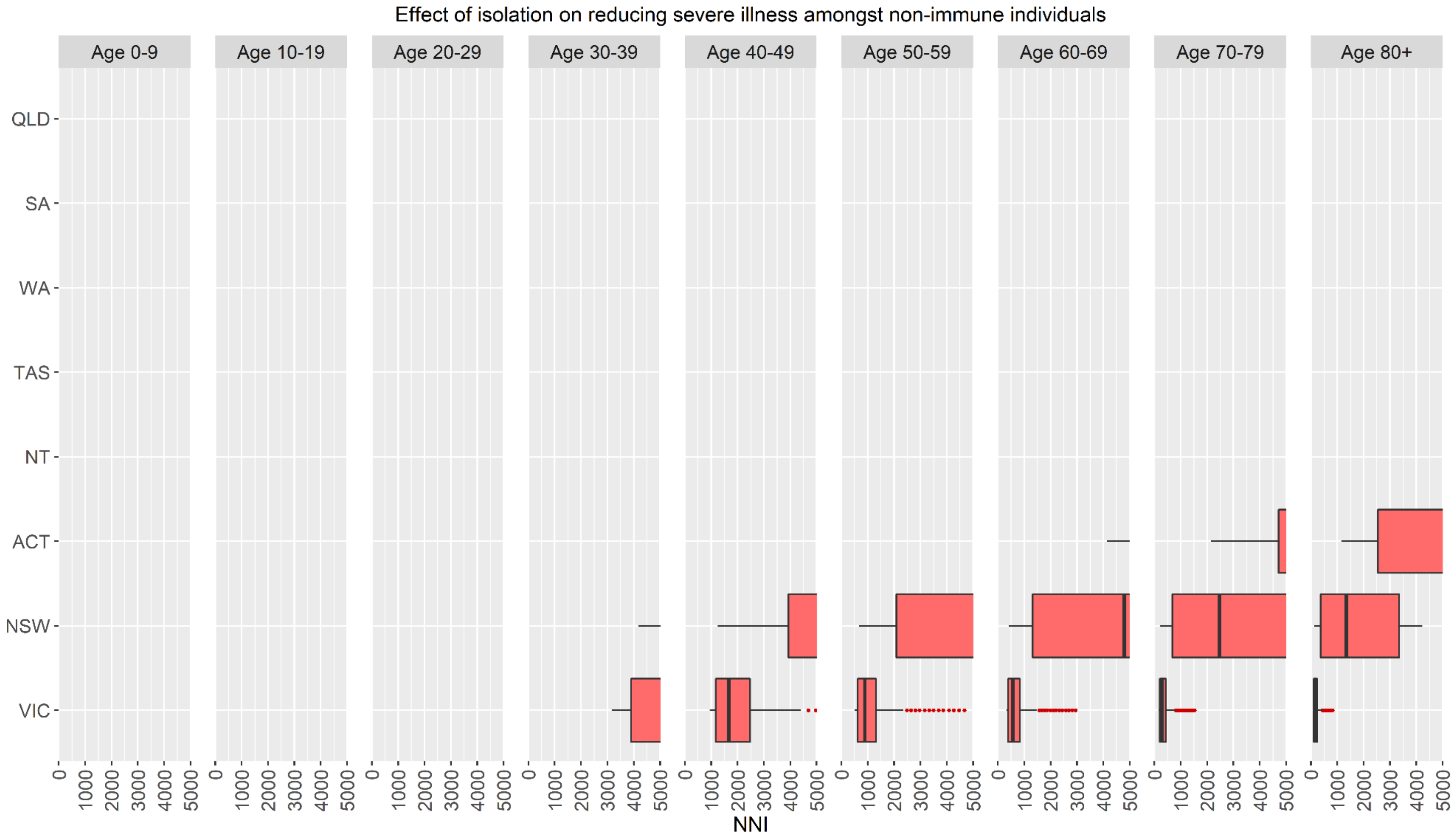

A region/age-group with no box-and-whisker plot has an NNI > 5,000 indicating a very low absolute risk reduction. ACT=Australian Capital Territory, NSW=New South Wales, NT=Northern Territory, QLD=Queensland, SA=South Australia, TAS=Tasmania, VIC=Victoria, WA=Western Australia.

**Figure S18.** Australia: NNIs for critical illness.

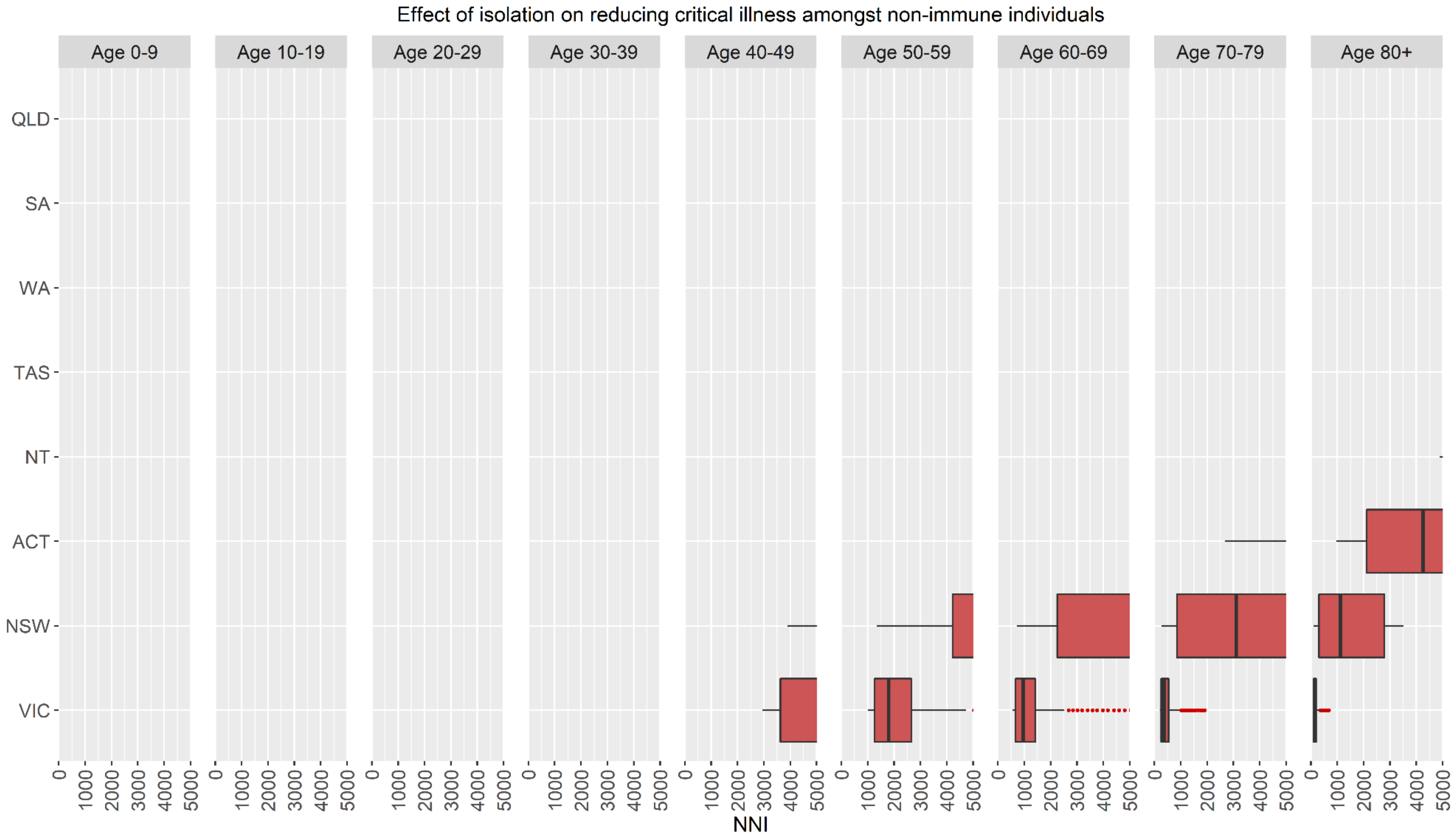

A region/age-group with no box-and-whisker plot has an NNI > 5,000 indicating a very low absolute risk reduction. ACT=Australian Capital Territory, NSW=New South Wales, NT=Northern Territory, QLD=Queensland, SA=South Australia, TAS=Tasmania, VIC=Victoria, WA=Western Australia.

**Figure S19.** US Northeast: NNIs for transmission in non-household settings.

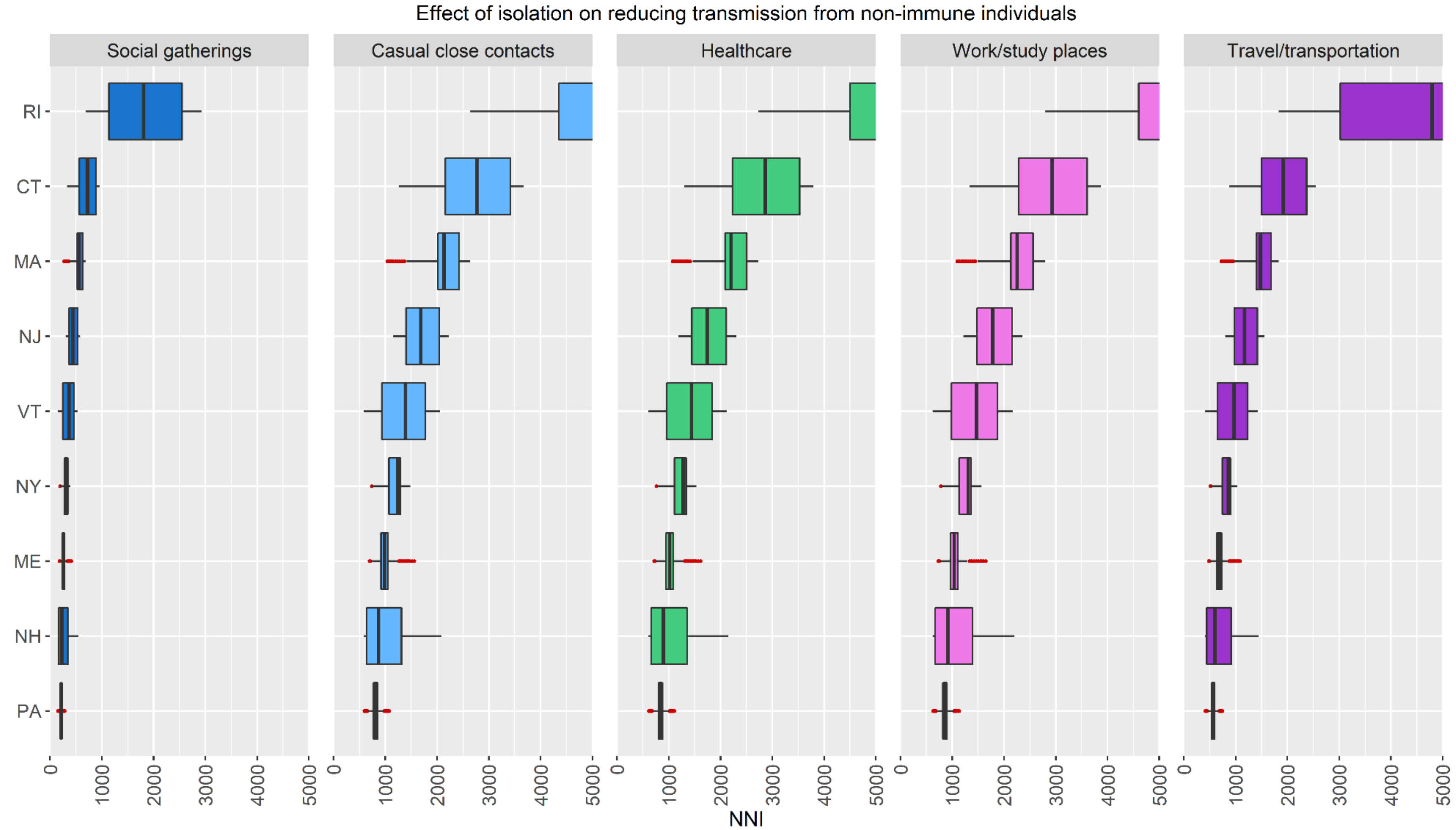

A region/setting with no box-and-whisker plot has an NNI > 5,000 indicating a very low absolute risk reduction. CT=Connecticut, ME=Maine, MA=Massachusetts, NH=New Hampshire, NJ=New Jersey, NY=New York, PA=Pennsylvania, RI=Rhode Island, VT=Vermont.

**Figure S20.** US Northeast: NNIs for severe illness.

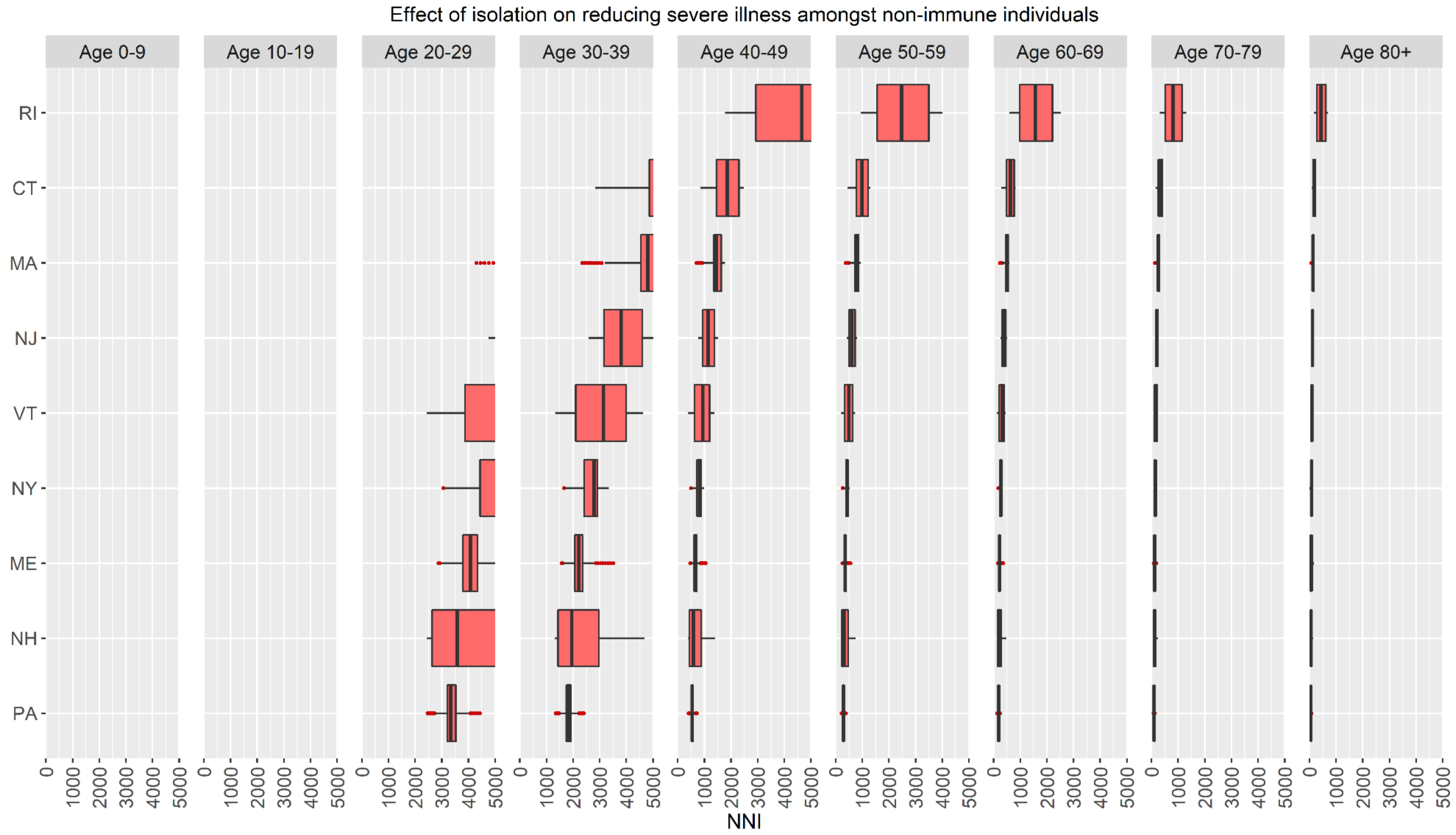

A region/age-group with no box-and-whisker plot has an NNI > 5,000 indicating a very low absolute risk reduction. CT=Connecticut, ME=Maine, MA=Massachusetts, NH=New Hampshire, NJ=New Jersey, NY=New York, PA=Pennsylvania, RI=Rhode Island, VT=Vermont.

**Figure S21.** US Northeast: NNIs for critical illness.

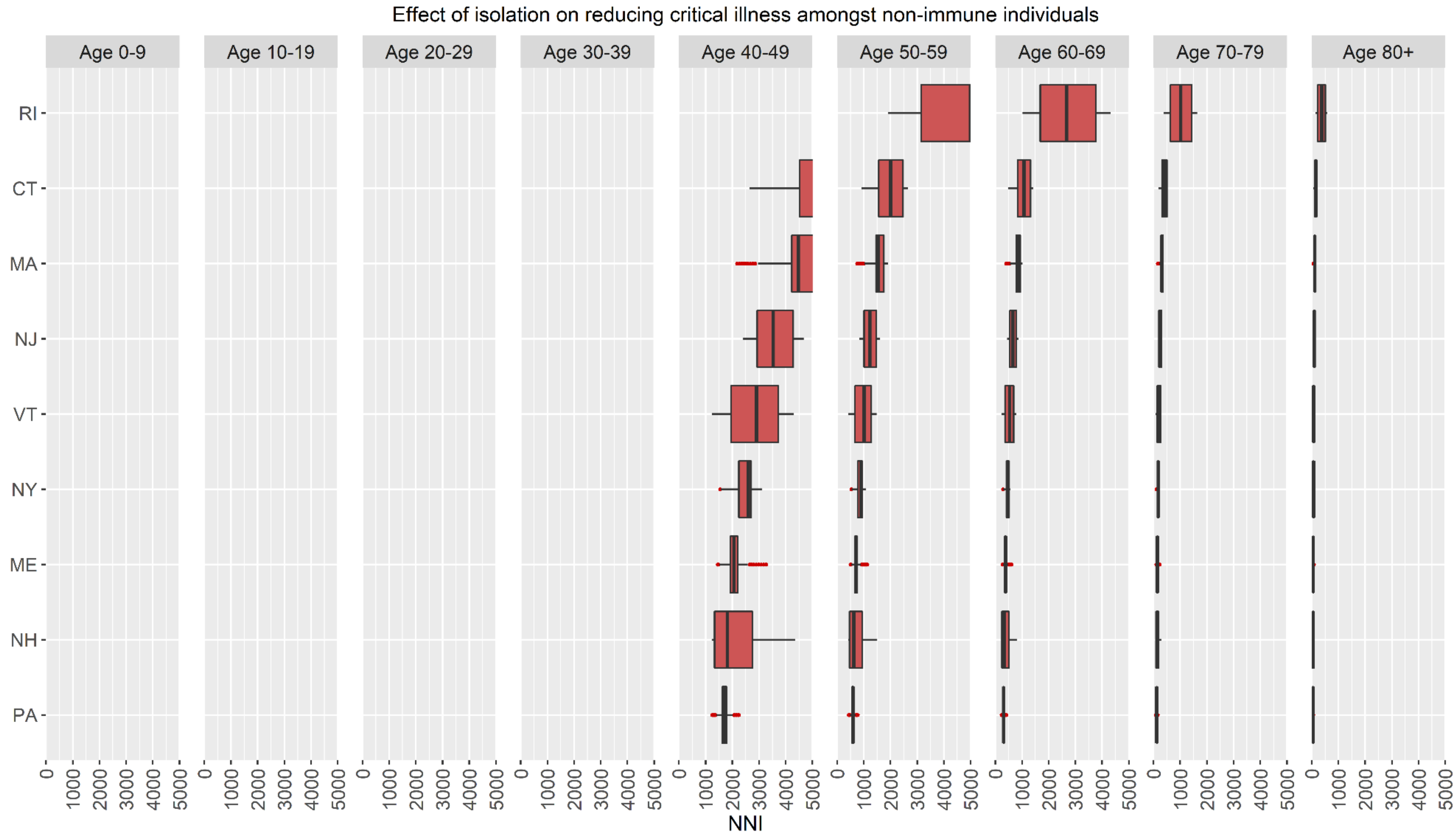

A region/age-group with no box-and-whisker plot has an NNI > 5,000 indicating a very low absolute risk reduction. CT=Connecticut, ME=Maine, MA=Massachusetts, NH=New Hampshire, NJ=New Jersey, NY=New York, PA=Pennsylvania, RI=Rhode Island, VT=Vermont.

**Figure S22.** US Midwest: NNIs for transmission in non-household settings.

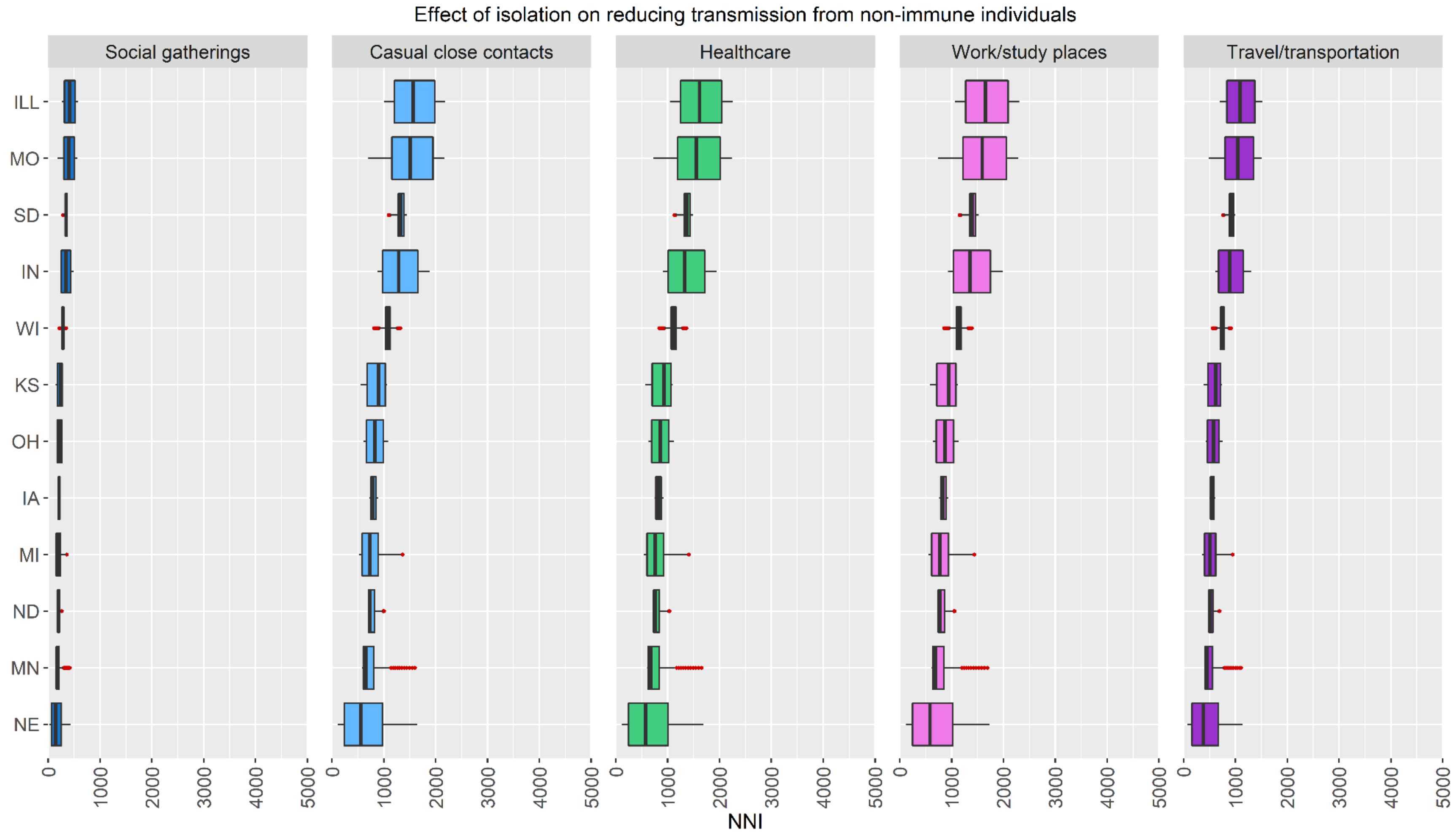

A region/setting with no box-and-whisker plot has an NNI > 5,000 indicating a very low absolute risk reduction. ILL=Illinois, IN=Indiana, IA=Iowa, KS=Kansas, MI=Michigan, MN=Minnesota, MO=Missouri, NE=Nebraska, ND=North Dakota, OH=Ohio, SD=South Dakota, WI=Wisconsin.

**Figure S23.** US Midwest: NNIs for severe illness.

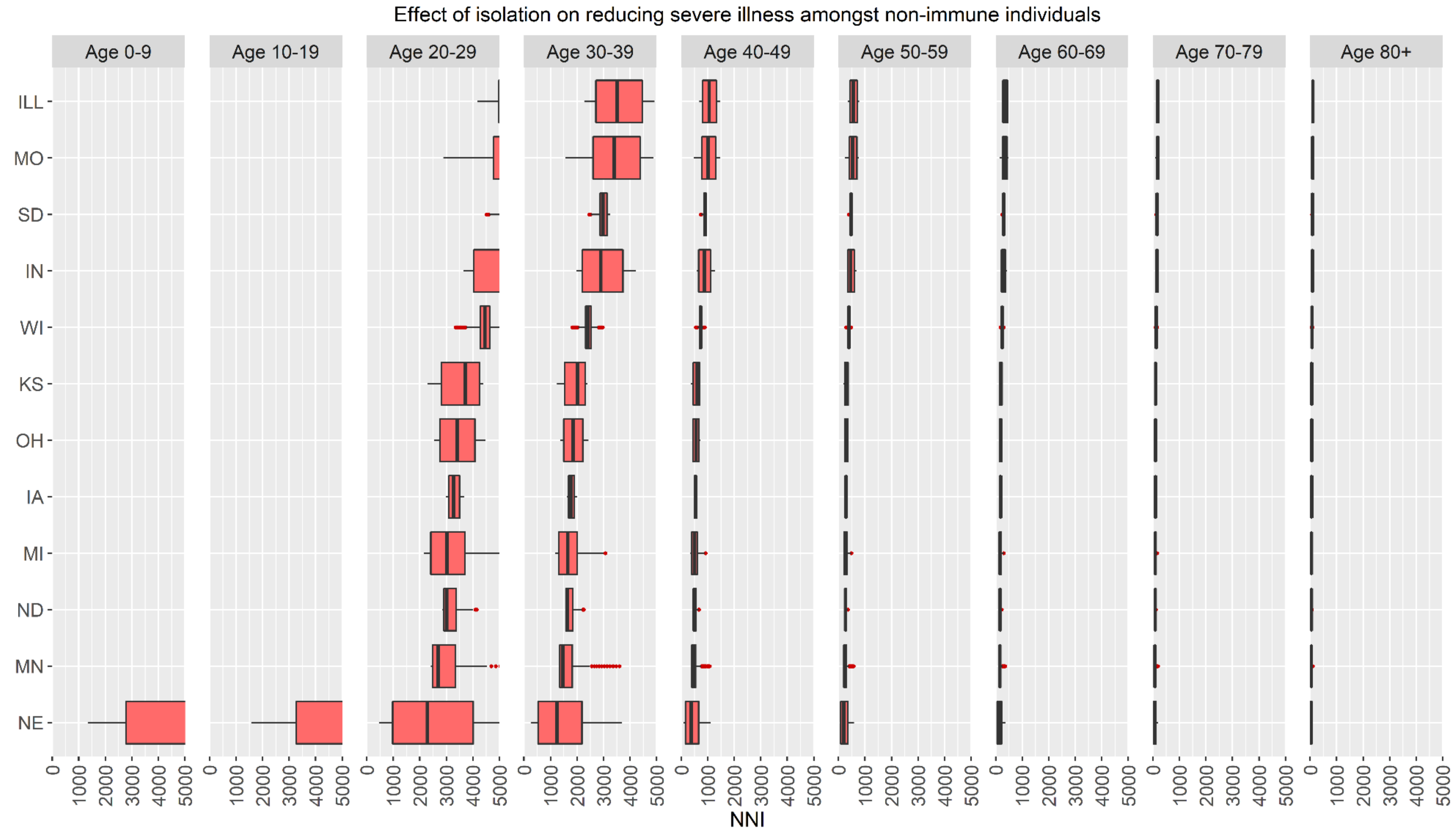

A region/age-group with no box-and-whisker plot has an NNI > 5,000 indicating a very low absolute risk reduction. ILL=Illinois, IN=Indiana, IA=Iowa, KS=Kansas, MI=Michigan, MN=Minnesota, MO=Missouri, NE=Nebraska, ND=North Dakota, OH=Ohio, SD=South Dakota, WI=Wisconsin.

**Figure S24.** US Midwest: NNIs for critical illness.

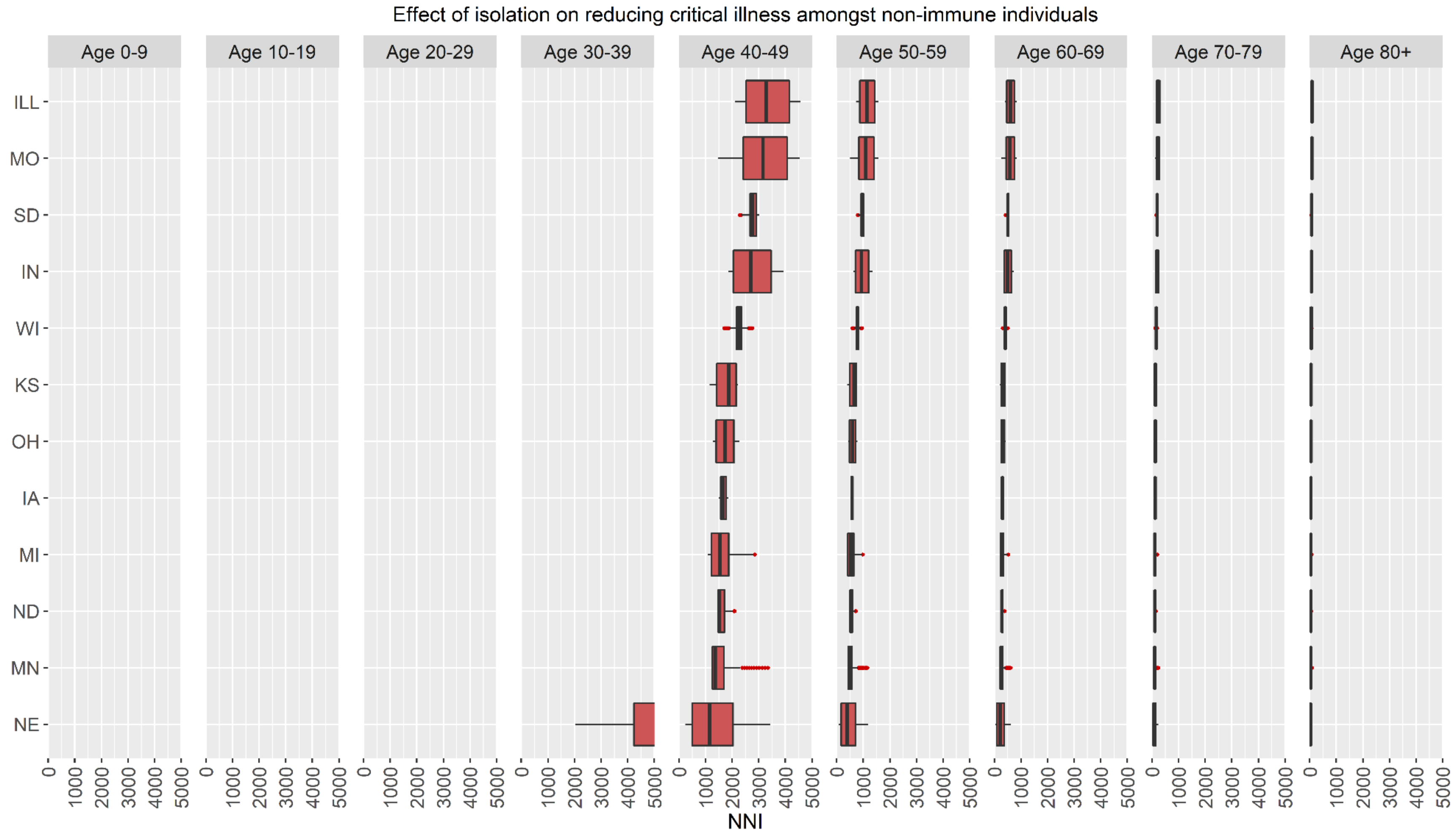

A region/age-group with no box-and-whisker plot has an NNI > 5,000 indicating a very low absolute risk reduction. ILL=Illinois, IN=Indiana, IA=Iowa, KS=Kansas, MI=Michigan, MN=Minnesota, MO=Missouri, NE=Nebraska, ND=North Dakota, OH=Ohio, SD=South Dakota, WI=Wisconsin.

**Figure S25.** US South: NNIs for transmission in non-household settings.

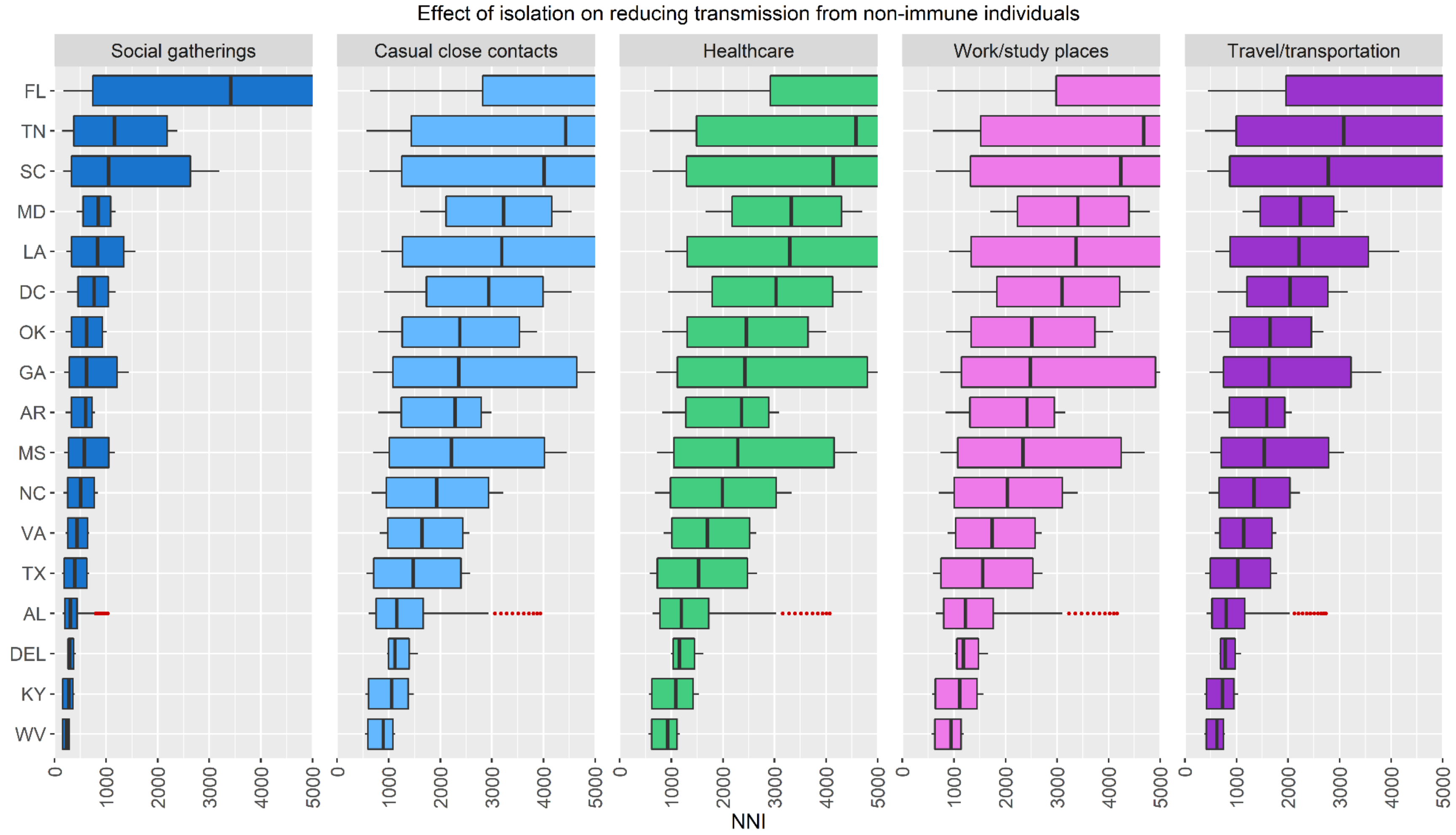

A region/setting with no box-and-whisker plot has an NNI > 5,000 indicating a very low absolute risk reduction. AL=Alabama, AR=Arkansas, DEL=Delaware, DC=District of Columbia, FL=Florida, GA=Georgia, KY=Kentucky, LA=Louisiana, MD=Maryland, MS=Mississippi, NC=North Carolina, OK=Oklahoma, SC=South Carolina, TN=Tennessee, TX=Texas, VA=Virginia, WV=West Virginia.

**Figure S26.** US South: NNIs for severe illness.

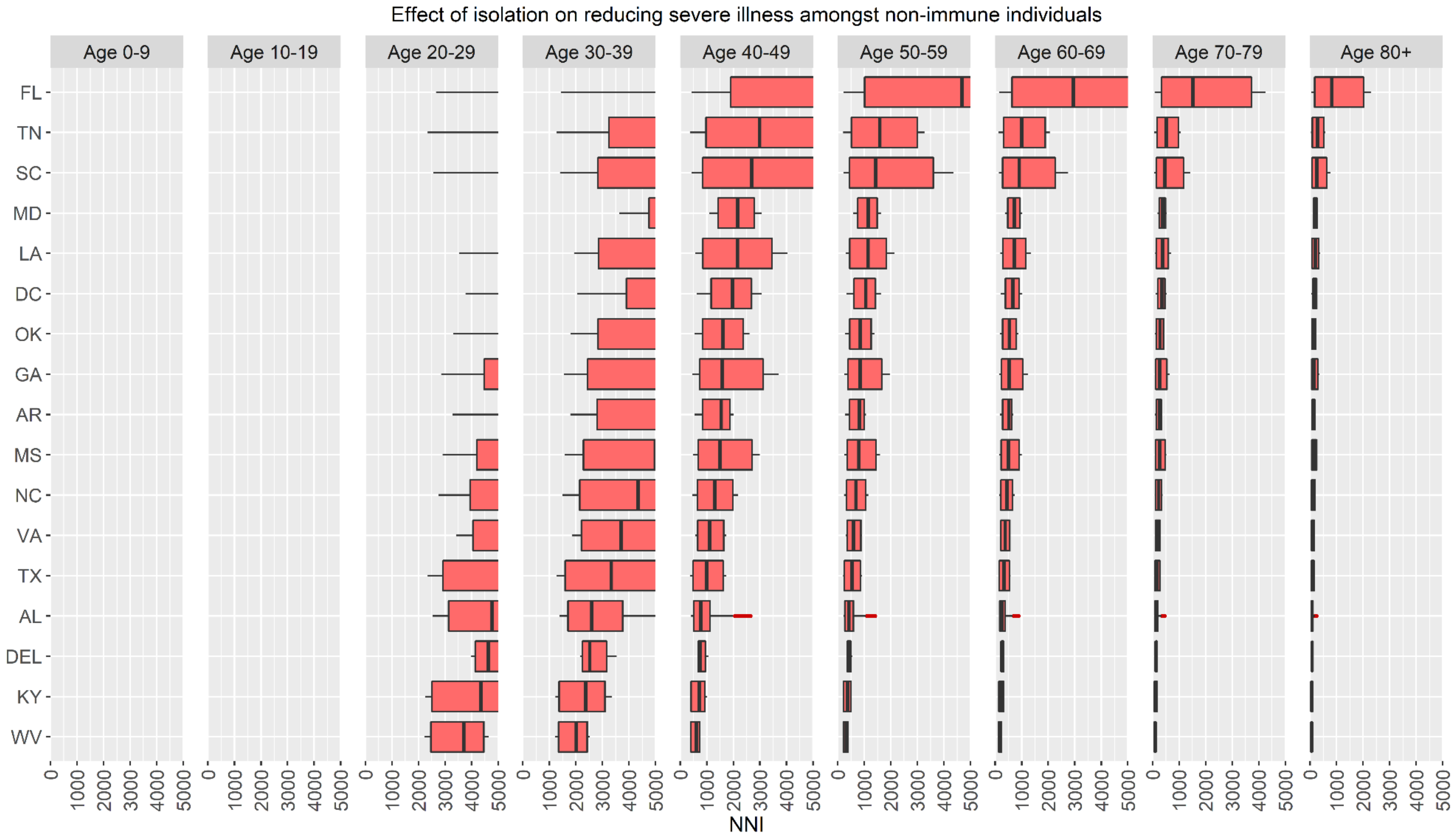

A region/age-group with no box-and-whisker plot has an NNI > 5,000 indicating a very low absolute risk reduction. AL=Alabama, AR=Arkansas, DEL=Delaware, DC=District of Columbia, FL=Florida, GA=Georgia, KY=Kentucky, LA=Louisiana, MD=Maryland, MS=Mississippi, NC=North Carolina, OK=Oklahoma, SC=South Carolina, TN=Tennessee, TX=Texas, VA=Virginia, WV=West Virginia.

**Figure S27.** US South: NNIs for critical illness.

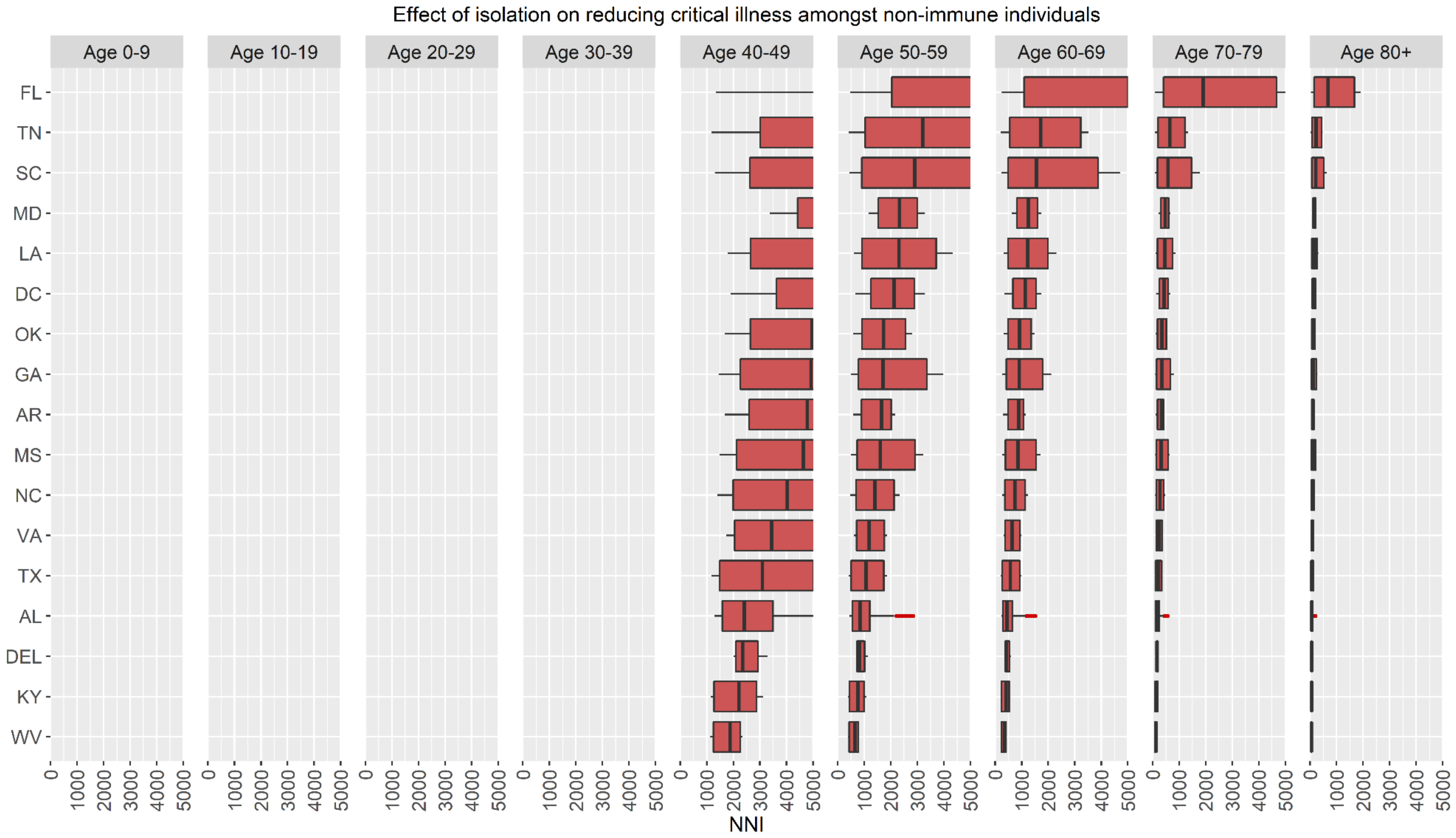

A region/age-group with no box-and-whisker plot has an NNI > 5,000 indicating a very low absolute risk reduction. AL=Alabama, AR=Arkansas, DEL=Delaware, DC=District of Columbia, FL=Florida, GA=Georgia, KY=Kentucky, LA=Louisiana, MD=Maryland, MS=Mississippi, NC=North Carolina, OK=Oklahoma, SC=South Carolina, TN=Tennessee, TX=Texas, VA=Virginia, WV=West Virginia.

**Figure S28.** US West: NNIs for transmission in non-household settings.

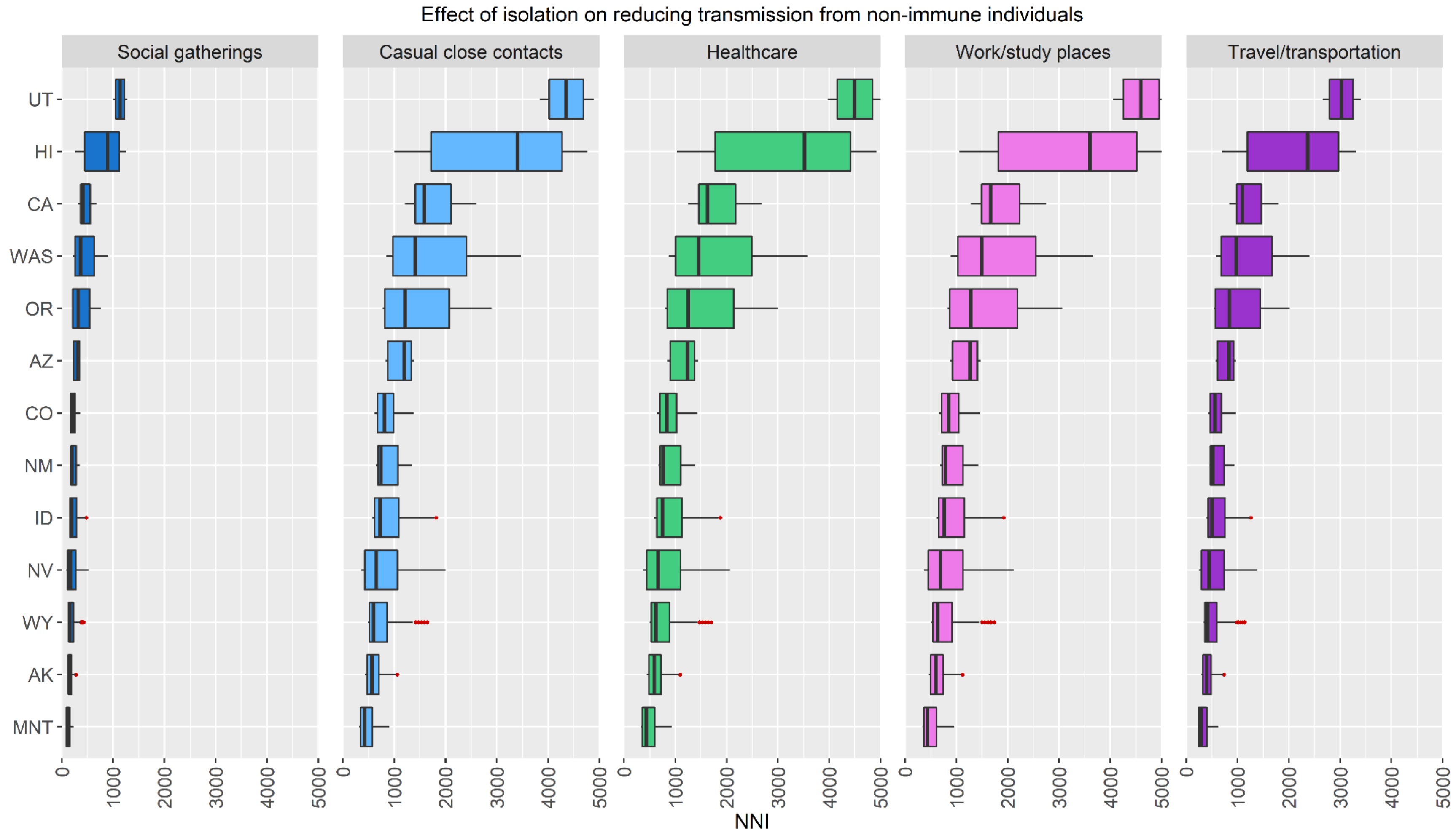

A region/setting with no box-and-whisker plot has an NNI > 5,000 indicating a very low absolute risk reduction. AK=Alaska, AZ=Arizona, CA=California, CO=Colorado, HI=Hawaii, ID=Idaho, MNT=Montana, NV=Nevada, NM=New Mexico, OR=Oregon, UT=Utah, WAS=Washington, WY=Wyoming.

**Figure S29.** US West: NNIs for severe illness.

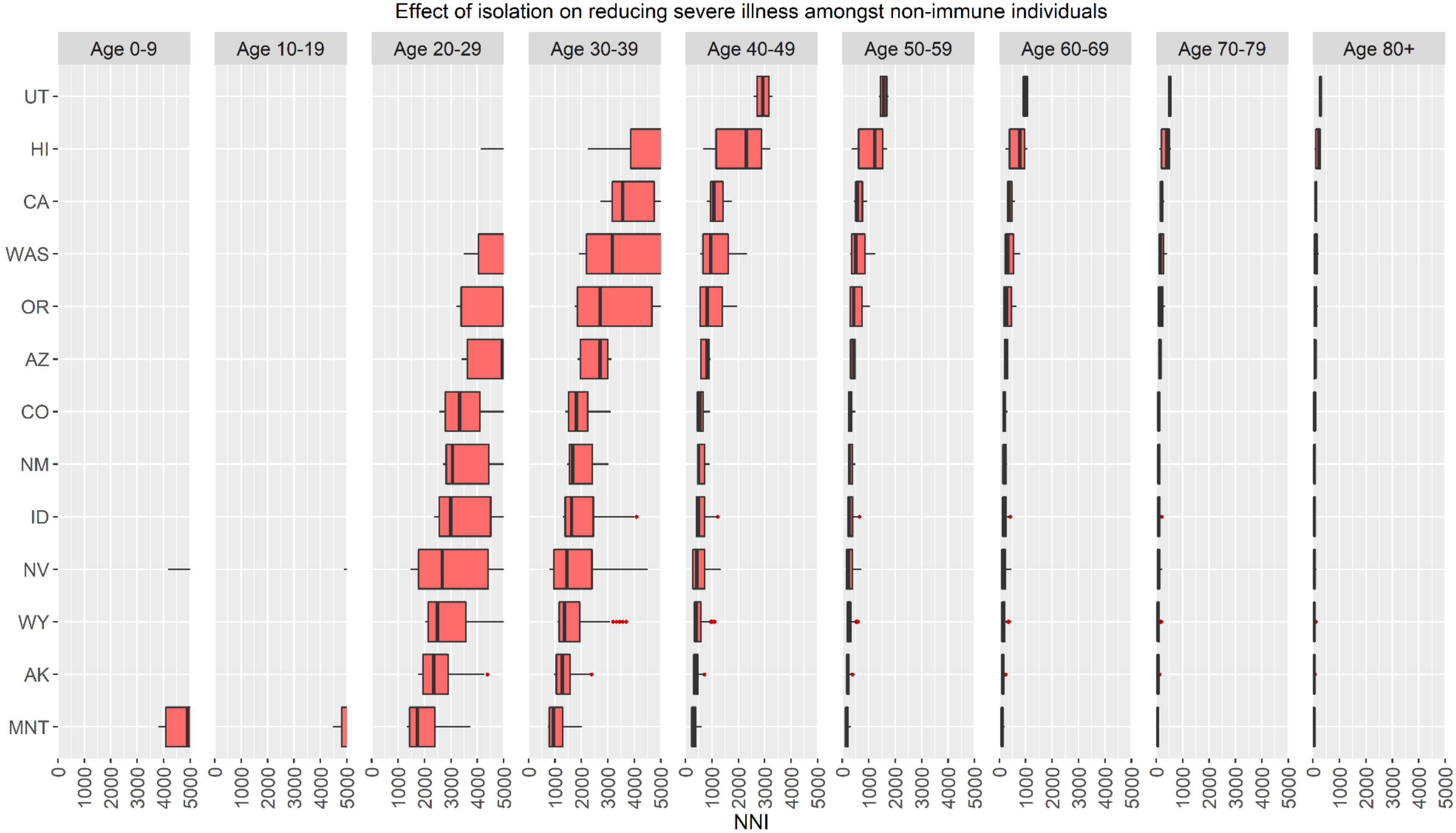

A region/age-group with no box-and-whisker plot has an NNI > 5,000 indicating a very low absolute risk reduction. AK=Alaska, AZ=Arizona, CA=California, CO=Colorado, HI=Hawaii, ID=Idaho, MNT=Montana, NV=Nevada, NM=New Mexico, OR=Oregon, UT=Utah, WAS=Washington, WY=Wyoming.

**Figure S30.** US West: NNIs for critical illness.

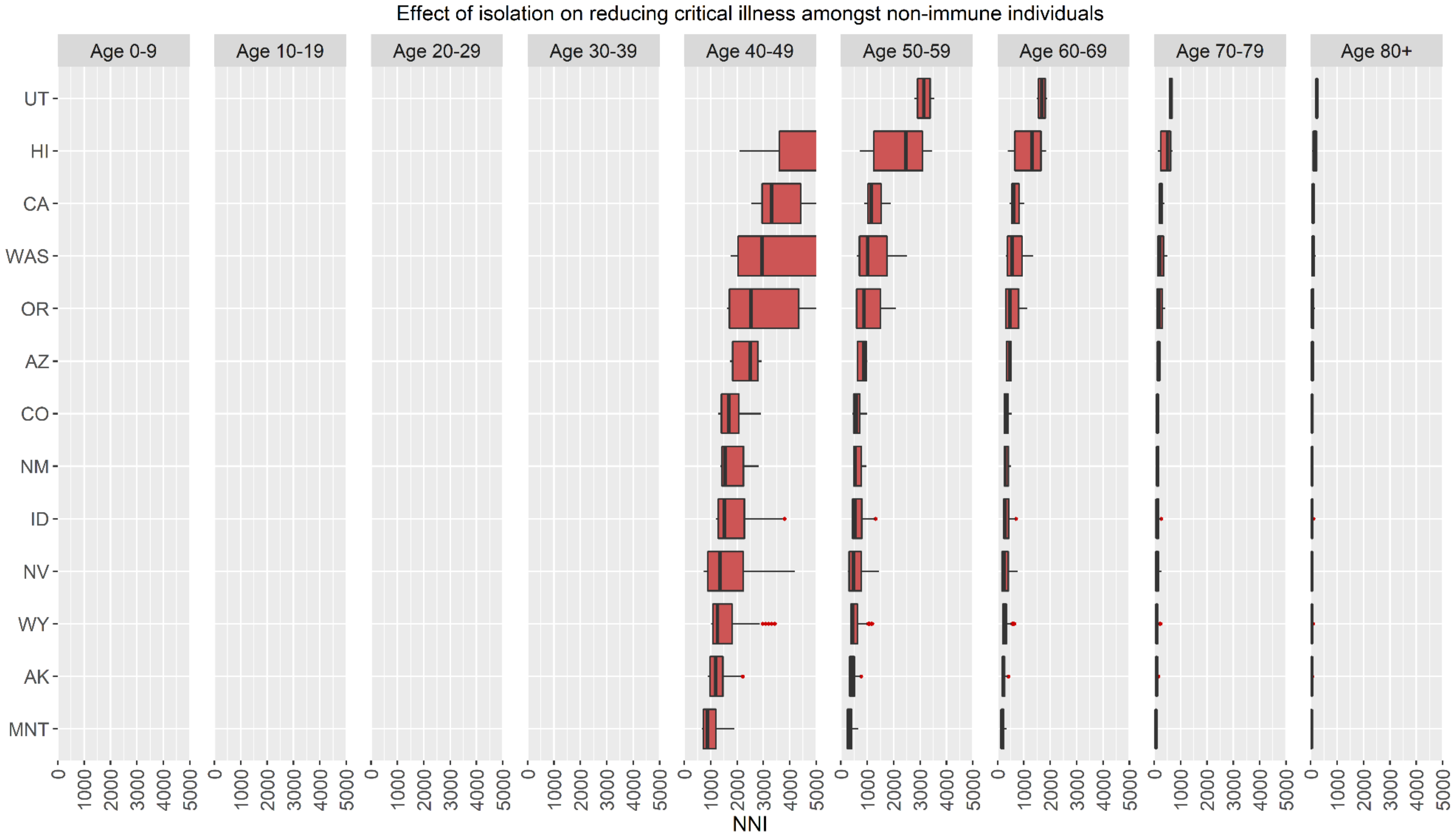

A region/age-group with no box-and-whisker plot has an NNI > 5,000 indicating a very low absolute risk reduction. AK=Alaska, AZ=Arizona, CA=California, CO=Colorado, HI=Hawaii, ID=Idaho, MNT=Montana, NV=Nevada, NM=New Mexico, OR=Oregon, UT=Utah, WAS=Washington, WY=Wyoming.
